# Quercetin activates the SIRT6-Nrf2 axis during oxidative stress, modulating ageing-associated markers in healthy men

**DOI:** 10.1101/2025.10.31.25338366

**Authors:** Ciara G. Juan, Nina Šimunić-Briški, Fabiana Volpe-Zanutto, Kyle Matchett, Gareth W. Davison

## Abstract

Quercetin is a dietary flavonoid with antioxidant, anti-inflammatory, and senolytic properties, yet its effects on exercise-induced DNA damage responses in healthy humans are unclear. We conducted a double-blind, placebo-controlled crossover trial (n = 13, middle-aged men) combining acute high-intensity interval exercise (HIIE) with 21-day quercetin supplementation (1000 mg/day). We quantified plasma quercetin and nuclear localisation, DNA repair and sirtuin gene expression, oxidative DNA damage (single and double-strand breaks, oxidised purines), and plasma/IgG glycomics including a preliminary GlycanAge index. Quercetin increased in plasma and nucleus post-exercise (p < 0.001; p < 0.05). Compared with placebo, quercetin increased post-exercise SIRT6-quercetin nuclear colocalisation (p < 0.001 at all timepoints) and Nrf2 nuclear translocation (p < 0.001). In placebo, *SIRT1, PARP1* and *RAD51* expression increased post-exercise, whereas *SIRT6* and *OGG1* decreased; this pattern was reversed with quercetin (all p < 0.05). Oxidative DNA damage markers were reduced with quercetin versus placebo (p < 0.001). Exercise-induced increases in plasma galactosylation and decreases in agalactosylation were observed in placebo but not with quercetin (all p < 0.05); GlycanAge showed non-significant increases post-exercise in placebo and a non-significant decrease following 21-day quercetin supplementation. In summary, short-term quercetin activates a SIRT6-Nrf2-linked response during HIIE, sustains DNA repair capacity, and stabilises inflammatory glycomic dynamics, consistent with a shift toward energetically economical, chromatin-associated repair. By providing evidence that quercetin biases DNA repair pathway usage under physiological oxidative stress, this work advances a framework of repair efficiency and energetic economy in human genome maintenance.

**Graphical abstract:** 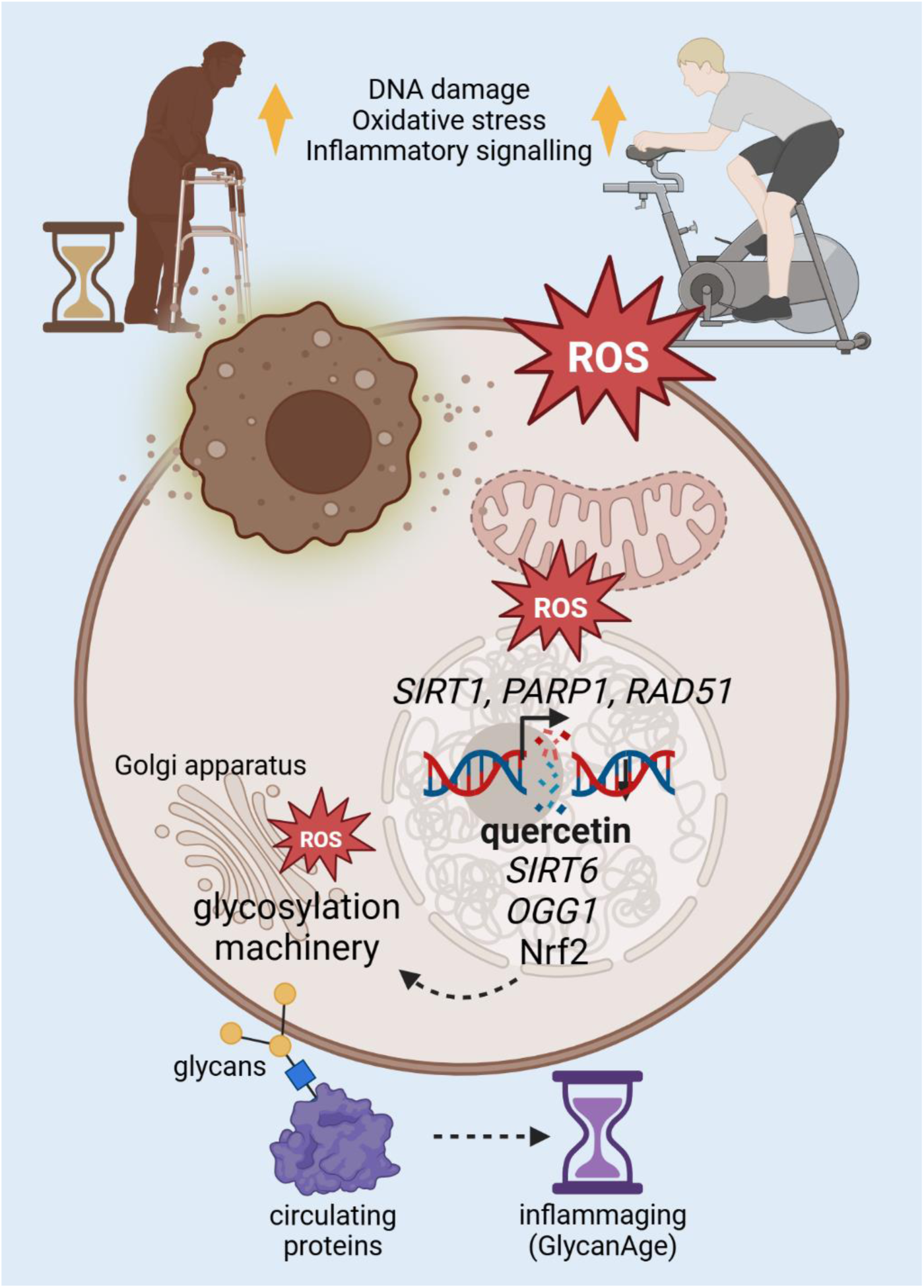

Ageing and high-intensity interval exercise (HIIE) engage overlapping oxidative stress, DNA damage, and inflammatory signalling pathways, but with distinct temporal profiles. Acute HIIE induces reactive oxygen species (ROS) and increases peripheral blood mononuclear cell (PBMC) expression of stress-responsive DNA damage response genes PARP1, SIRT1, and RAD51, whereas SIRT6, OGG1, and nuclear Nrf2 are selectively increased with quercetin supplementation. Exercise-induced plasma N-glycosylation remodelling—characterised by increased galactosylation and reduced agalactosylation—was observed after HIIE but was abolished by quercetin supplementation, indicating stabilisation of glycosylation dynamics under reduced oxidative signalling. GlycanAge (an inflammaging index) exhibited only non-significant acute changes but may require longer-term supplementation to shift measurably. Dashed arrows indicate inferred or integrative relationships.

## Introduction

Oxidative stress arises when the generation of reactive oxygen and nitrogen species (RONS) surpasses the capacity of endogenous antioxidant defences to neutralise them. This imbalance leads to the oxidation of lipids, proteins, and nucleic acids, with DNA being a particularly susceptible target due to its constant exposure to metabolic by-products and environmental insults. Oxidative DNA lesions encompass base modifications such as 8-oxo-7,8-dihydroguanine (8-oxoG, commonly quantified as 8-oxodG), abasic (AP) sites, and both single and double-strand breaks (DSB), which can compromise genomic integrity if not efficiently repaired. In mammalian cells, several tightly regulated repair pathways maintain DNA stability: the base excision repair (BER) pathway recognises and excises oxidised bases and single-strand breaks through the coordinated activity of DNA glycosylases such as OGG1, XRCC1, and PARP1, while DSB are resolved through either homologous recombination (HR) involving RAD51, or non-homologous end joining (NHEJ), all of which are regulated by SIRT1 and SIRT6. With advancing age, redox imbalance, chronic inflammation, and reduced NAD⁺ bioavailability attenuate the efficiency of DNA repair, resulting in DNA damage accumulation, genomic instability, and tissue dysfunction. Enhancing DNA repair capacity and redox homeostasis is therefore a central focus of emerging geroprotective interventions aimed at promoting healthy ageing.

Acute high-intensity interval exercise (HIIE) is a non-pharmacological, ethically acceptable, physiologically relevant model of oxidative stress and DNA damage (Tryfidou et al., 2020). Meanwhile, repeated moderate to high-intensity exercise training induces transient oxidative stress that elicits adaptive upregulation of endogenous antioxidant and DNA repair systems (Soares et al., 2015). This hormetic response involves activation of redox-sensitive transcription factors such as Nrf2, FOXO, and PGC-1α, leading to increased expression of antioxidant enzymes, more efficient DNA repair, and reduced genomic damage (Denham et al., 2015; Powers et al., 2016). Mechanistically, RONS generated during muscle contraction—primarily H₂O₂—serve as critical second messengers that activate the NF-κB and MAPK cascades, two central pathways in redox-regulated gene expression (Ji, 2007). RONS transiently oxidise cysteine residues on protein tyrosine phosphatases, sustaining kinase phosphorylation and activating the IκB kinase (IKK) complex, which liberates NF-κB to bind promoter regions of antioxidant genes such as MnSOD and GPX. Concurrently, exercise stimulates phosphorylation of ERK1/2, JNK, and p38MAPK, which phosphorylate downstream transcription factors and reinforce NF-κB-dependent antioxidant gene transcription (Ji, 2007). This cross-talk between MAPK and NF-κB ensures integrated regulation of antioxidant enzymes, nitric oxide synthase, and mitochondrial biogenesis via PGC-1α, contributing to the long-term enhancement of oxidative stress tolerance (Ji, 2007).

From a nutritional perspective, quercetin, a polyphenolic flavonol, possesses complementary bioactivities that intersect with exercise-induced adaptive pathways. Beyond its well-characterised senolytic and radical-scavenging capacity, quercetin exerts pleiotropic effects on key regulators of metabolism, DNA repair, and inflammation. It activates AMPK and sirtuin signalling (particularly SIRT1 and SIRT6), enhances Nrf2-Keap1 pathway activation, and modulates NF-κB-mediated inflammatory responses (Abd El-Emam et al., 2023; Lu et al., 2023). Structural and molecular docking studies have demonstrated direct binding of quercetin to SIRT6 and AMPK catalytic domains, suggesting a potential for allosteric activation of pathways integral to chromatin stability, BER efficiency, and NAD⁺ homeostasis (Lu et al., 2023; You et al., 2019). In animal models, quercetin has been shown to reduce oxidative DNA damage, upregulate OGG1 and XRCC1, and attenuate senescence-associated secretory phenotypes, supporting its classification as a candidate senolytic and DNA repair modulator (Bujarrabal-Dueso et al., 2025; Darband et al., 2020).

Despite robust evidence for exercise and quercetin as individual modulators of redox and repair pathways, their combined effects on oxidative DNA damage and repair in humans remain poorly characterised. Understanding whether quercetin can augment or synergise with exercise-induced DNA damage response is critical for developing integrative lifestyle and nutritional strategies to mitigate age-related genomic instability. The present study was therefore designed to examine the effects of HIIE and quercetin supplementation—individually and in combination—on molecular and biochemical markers of DNA damage and repair in healthy middle-aged men.

## Results

### Retention and Adherence

Participant retention was 54%, yielding a final analytical sample of n = 13. Despite attrition, the general linear model with a repeated-measures design indicated an observed power of 1.00 for the Time × Condition interaction, confirming adequate statistical power for the primary outcomes. Among retained participants, supplementation adherence was 100%, as reflected by linear increases in plasma quercetin aglycone levels in every participant, measured by UHPLC–MS/MS.

### Quercetin Levels and Uptake

Plasma quercetin aglycone levels increased in all active treatment timepoints (*p* < 0.05). Acute supplementation resulted in significantly higher plasma quercetin levels post-exercise compared to placebo (*p* < 0.001, Figure 1A). Meanwhile, 21-day supplementation resulted in significantly higher quercetin levels both at rest and post-exercise (*p* < 0.001 for both). In addition, nuclear quercetin increased post-exercise in both treatment conditions (all *p* < 0.05). Short-term supplementation increased the post-exercise nuclear uptake of quercetin relative to placebo (*p* < 0.05, Figure 1C).

**Figure 1.**
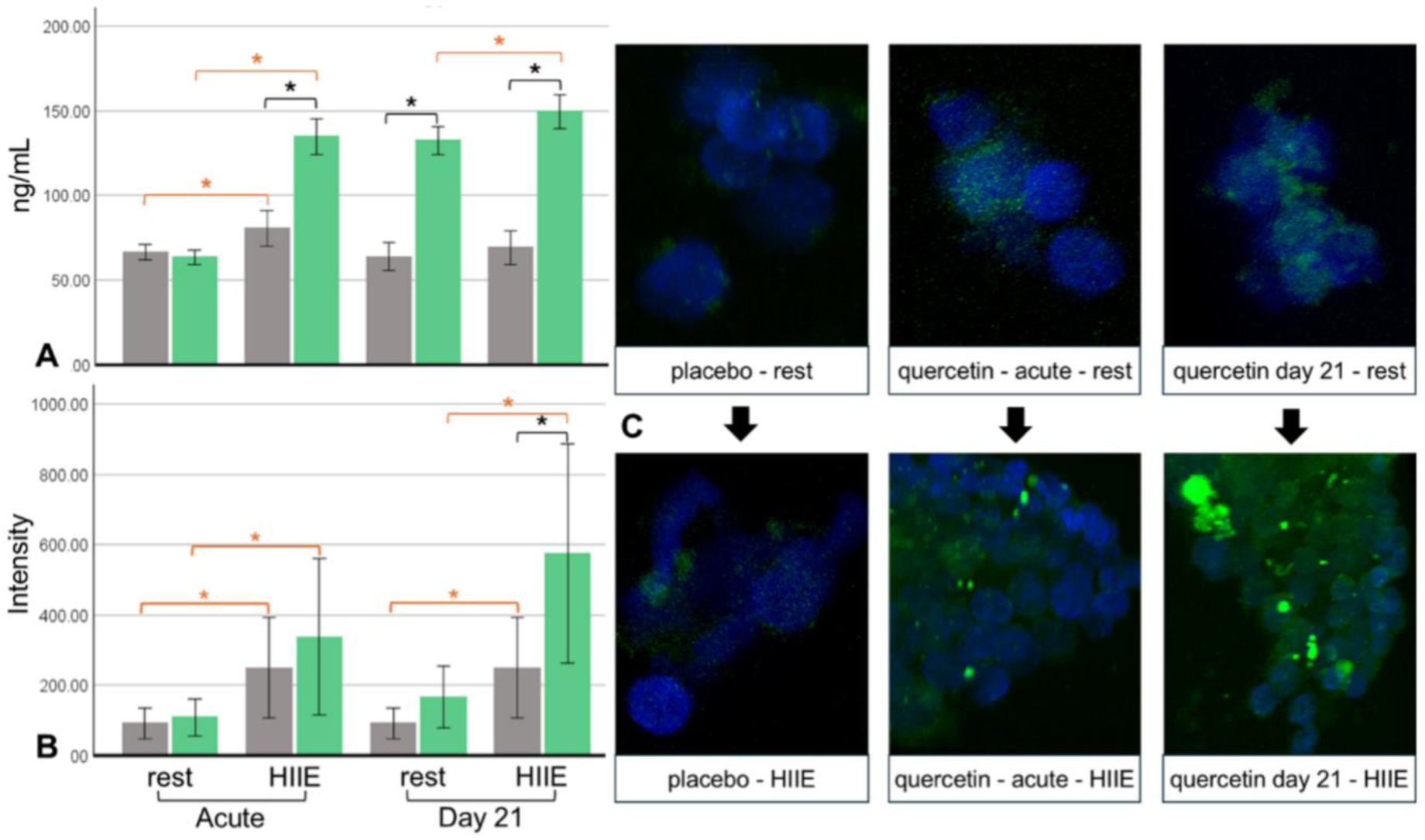
Quercetin levels and cellular uptake. A) Plasma quercetin aglycone levels (ng/ml) following acute and short-term phases of supplementation. B) Median fluorescence intensity of native, unstained quercetin in nucleus. C) Confocal microscopy of native, unstained quercetin. Black asterisk indicates significant between-treatment difference (quercetin vs placebo). Orange asterisk indicates significant within-subject difference (pre vs post-exercise). Green bars represent quercetin, grey bars represent placebo.

### DNA Damage

DNA strand breaks (comet assay) increased post-exercise in all exercise sessions and conditions (*p* < 0.001 for all timepoints, Figure 2A). Short-term supplementation significantly decreased the post-exercise strand break relative to placebo (*p* < 0.001). Similarly, purine base oxidation increased post-exercise in all exercise sessions and conditions (*p* < 0.001 for all timepoints, Figure 2B), with significantly lower oxidation post-exercise in quercetin compared to placebo after short-term supplementation (*p* < 0.001). DSB increased post-exercise in all exercise sessions and conditions (*p* < 0.001 for all timepoints, Figure 2D). Both acute and short-term supplementation resulted in lower DSB post-exercise compared to placebo (*p* < 0.001 for both).

**Figure 2.**
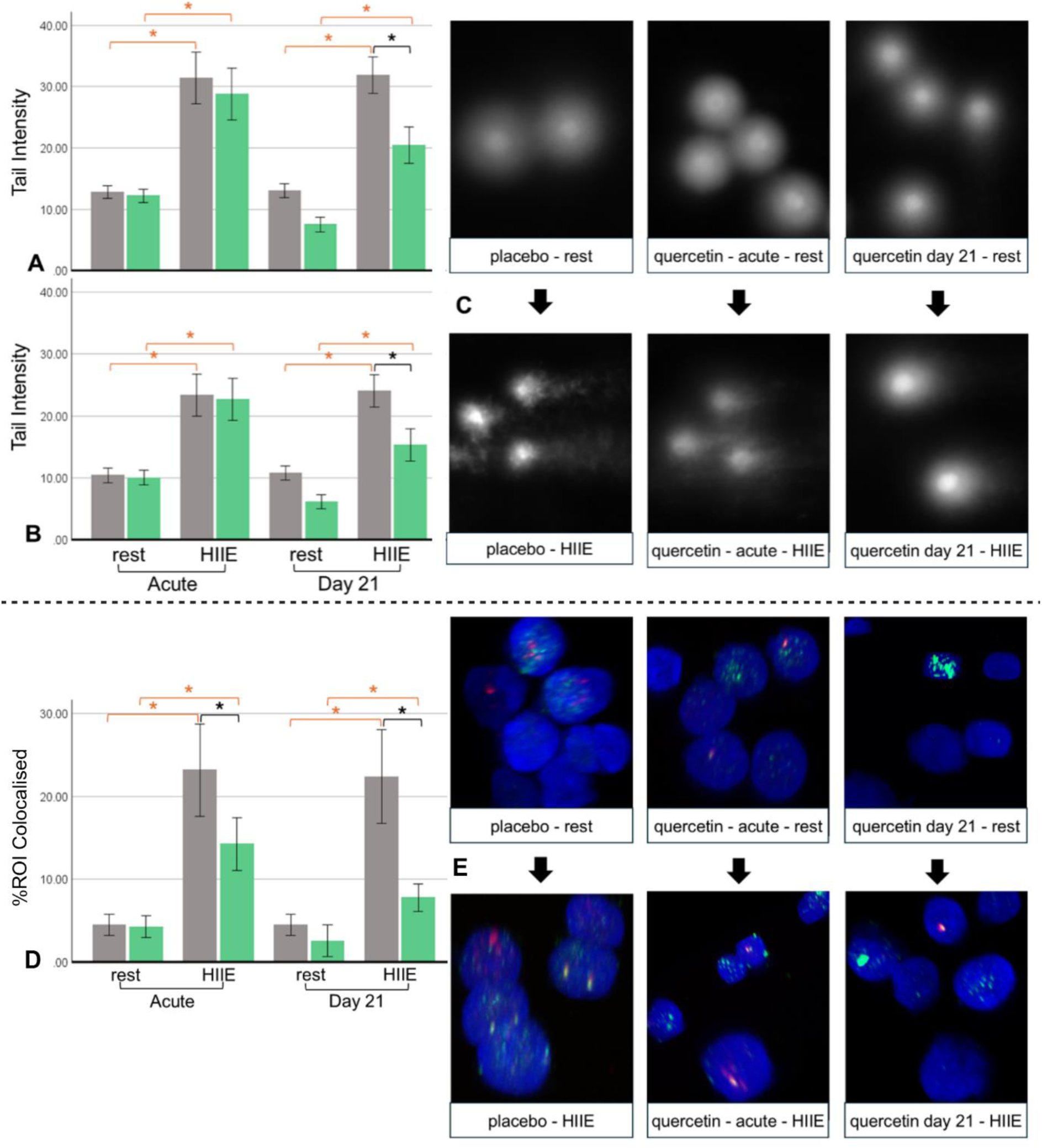
DNA damage markers. A) DNA strand breaks (tail intensity) quantified by single-cell gel electrophoresis. B) Oxidised purine bases (tail intensity). C) Fluorescence microscopy of DNA fragment migration. D) DNA double-strand breaks quantified by ƴH2AX and 53BP1 colocalisation (%ROI colocalised). E) Confocal microscopy of ƴH2AX and 53BP1 colocalisation showing white puncta from overlapping green (ƴH2AX) and red (53BP1). Black asterisk indicates significant between-treatment difference (quercetin vs placebo). Orange asterisk indicates significant within-subject difference (pre vs post-exercise). Green bars represent quercetin, grey bars represent placebo.

### Transcriptional DNA Repair

*SIRT1* expression significantly increased post-exercise in all placebo sessions (*p* < 0.05 for all, Figure 3A), but this effect was attenuated in quercetin. Meanwhile, *SIRT6* significantly decreased in all placebo sessions (*p* < 0.05 for all, Figure 3B), but increased post-exercise after short-term quercetin supplementation (*p* < 0.05). Similar to *SIRT1*, *PARP1* significantly increased post-exercise in all placebo sessions (*p* < 0.05 for all, Figure 3C), with no significant effects in quercetin. Meanwhile, *OGG1*, like *SIRT6*, significantly decreased post-exercise in all placebo sessions (*p* < 0.05 for all, Figure 3D), but significantly increased post-exercise in all quercetin sessions (*p* < 0.05 for all). Finally, *RAD51,* like *SIRT1* and *PARP1,* significantly increased post-exercise in all placebo sessions (*p* < 0.05 for all, Figure 3E), with no significant effects in quercetin. There were no significant changes in *XRCC1* expression.

**Figure 3.**
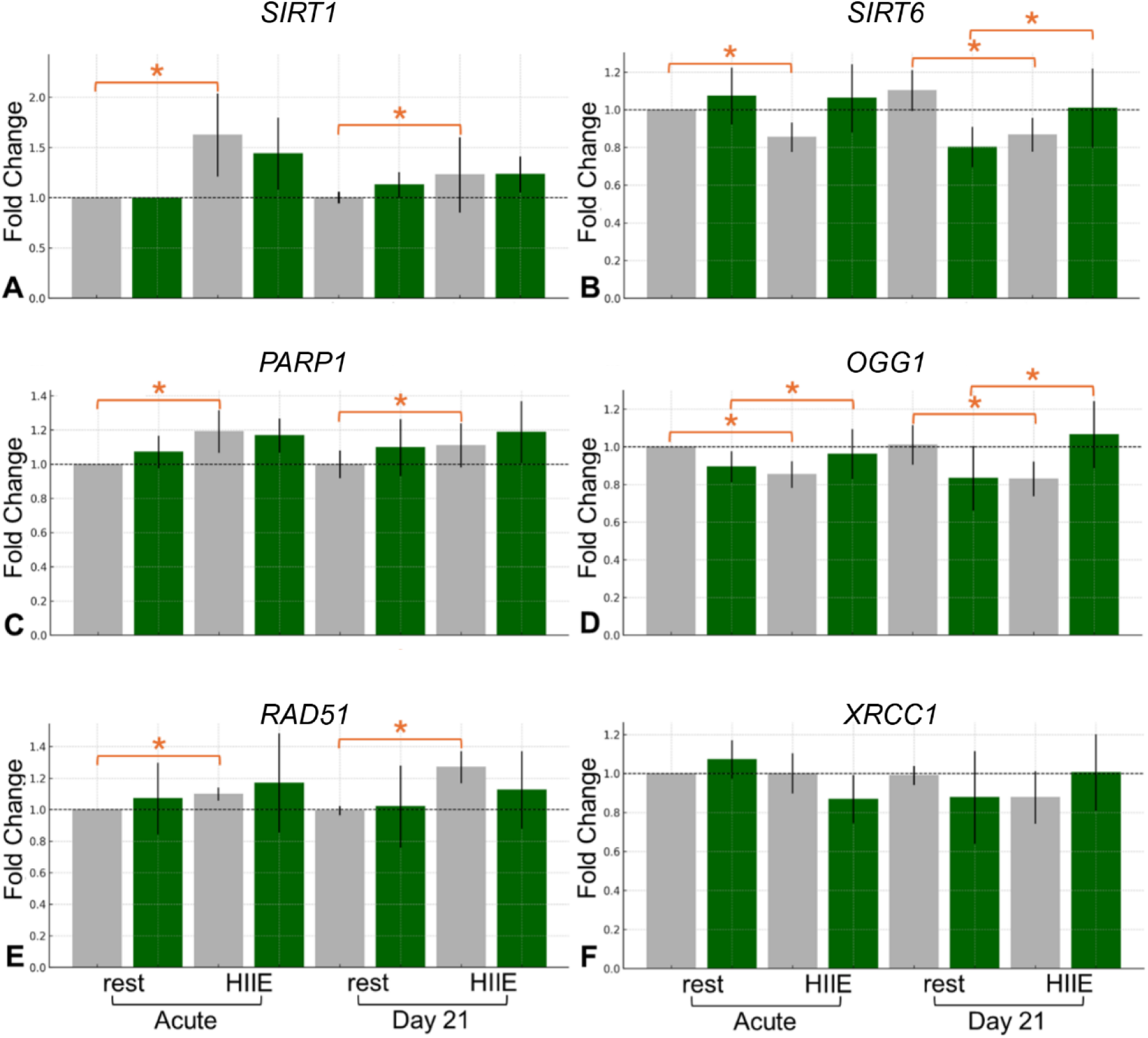
DNA repair gene expression. Fold changes in A) SIRT1; B) SIRT6; C) PARP1; D) OGG1; E) RAD51; and F) XRCC1. Orange asterisk indicates significant within-subject difference (pre vs post-exercise). Green bars represent quercetin, grey bars represent placebo.

Since SIRT6 has been shown to bind with quercetin, colocalisation of SIRT6 protein with quercetin was quantified using super-resolution 3D microscopy. Post-exercise colocalisation increased in all exercise sessions and conditions (*p* < 0.001 for all timepoints, Figure 4A). Both acute and short-term supplementation increased the post-exercise colocalisation relative to placebo (*p* < 0.001 for both). Short-term supplementation also increased resting colocalisation relative to placebo (*p* < 0.001).

**Figure 4.**
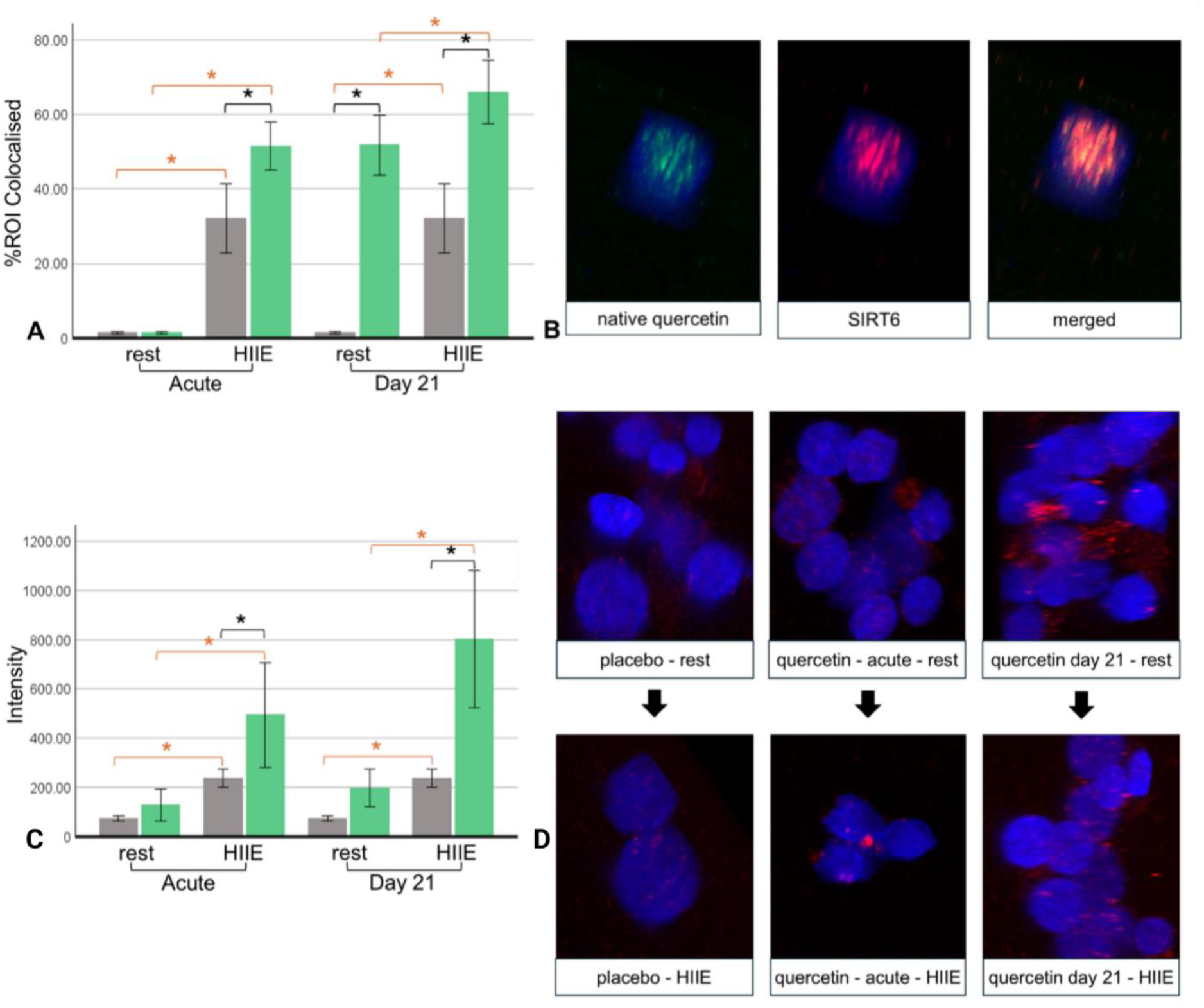
Localisation of SIRT6, quercetin, and Nrf2. A) SIRT6 colocalisation with native, unstained quercetin (%ROI colocalised). B) Confocal microscopy of colocalization. C) Nuclear Nrf2 (median intensity). D) Confocal microscopy of Nrf2 nuclear translocation. Black asterisk indicates significant between-treatment difference (quercetin vs placebo). Orange asterisk indicates significant within-subject difference (pre vs post-exercise). Green bars represent quercetin, grey bars represent placebo.

Since SIRT6 mono-ADP-ribosylation activity is essential for transcriptional activation of a subset of Nrf2 target genes under oxidative stress (Rezazadeh et al., 2019), Nrf2 was also imaged. Nuclear Nrf2 increased post-exercise in all exercise sessions and conditions (p < 0.001 for all timepoints, Figure 4C). Both acute and short-term quercetin supplementation increased nuclear Nrf2 post-exercise relative to placebo (p < 0.05 for both).

### Glycomic Profiling of IgG and Plasma Proteins

Plasma agalactosylation (G0), a pro-inflammatory marker, significantly decreased post-exercise in placebo (*p* < 0.05, Figure 5A), reflecting a reduction in pro-inflammatory agalactosylated structures and suggesting an acute anti-inflammatory response or post-exercise resolution phase. This acute response was not observed in the quercetin arm, suggesting that quercetin supplementation modulated the exercise-induced glycan shift. Plasma digalactosylation (G2) and total galactosylation (Gtotal), both anti-inflammatory markers, significantly increased post-exercise in placebo (both *p* < 0.05, Figure 5C and 5D), reflecting a transient anti-inflammatory glycomic shift. No significant changes were observed in IgG markers.

**Figure 5.**
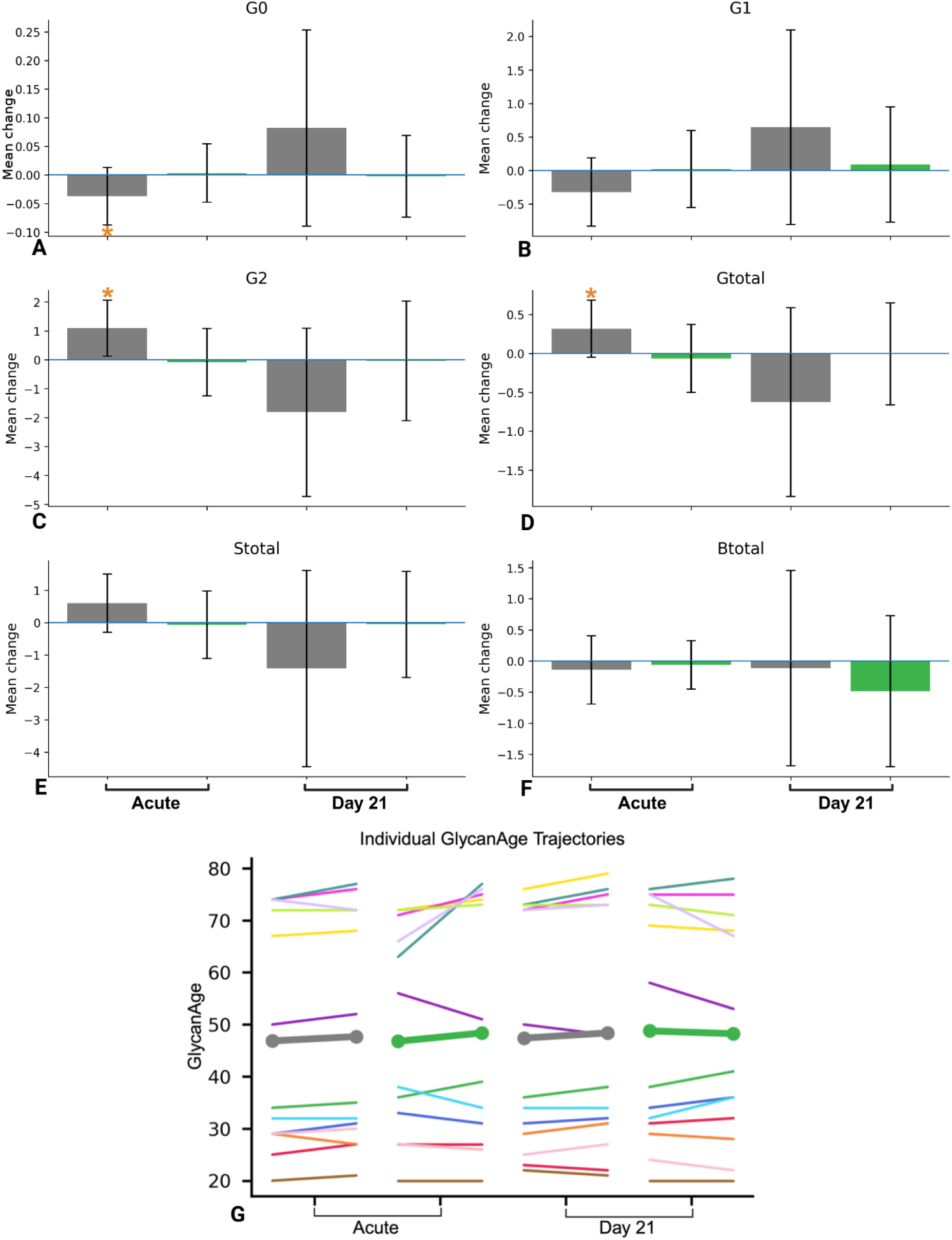
Plasma and IgG glycan changes. A) Plasma agalactosylation (G0); B) Plasma monogalactosylation (G1); C) Plasma digalactosylation (G2); D) Plasma total galactosylation (Gtotal); E) Plasma total sialylation (Stotal); F) Plasma total branching; and G) Individual IgG-based GlycanAge trajectories, with thin coloured lines representing within-individual changes for each participant, and bold green and grey lines with end-point markers denoting group mean GlycanAge. Orange asterisk indicates significant within-subject difference (pre vs post-exercise). Green lines and bars represent quercetin, grey lines and bars represent placebo.

### GlycanAge

Baseline GlycanAge at study entry ranged from 20 to 74 years across participants, with a mean of 47.8 ± 22.4 years, indicating marked inter-individual heterogeneity in glycan-derived biological ageing (Figure 5G). There was a non-significant trend towards increased GlycanAge index post-exercise in placebo, and a non-significant decrease post-exercise in quercetin after three-week supplementation.

## Discussion

Plasma quercetin aglycone concentrations increased at all timepoints in quercetin, confirming effective systemic delivery at the administered dose of 1,000 mg/day. Significant elevations were observed after the first dose and increased linearly following 21 days of continuous intake, consistent with previous reports demonstrating accumulation of quercetin and its metabolites in plasma over repeated dosing (Burak et al., 2017). Importantly, microscopy demonstrated nuclear localisation of native, unstained quercetin within PBMCs following acute HIIE across exercise sessions and treatment conditions. Under oxidative or inflammatory stress, deconjugation of quercetin glucuronides is enhanced, leading to increased intracellular accumulation of quercetin aglycone via local deconjugation and passive diffusion (Ishisaka et al., 2013).

Acute bouts of HIIE elicited increases in DNA damage, including strand breaks, oxidised purine bases, and DSB foci, consistent with prior work showing that intense aerobic exercise increases RONS production via mitochondrial electron transport chain activity, NADPH oxidase activation, and xanthine oxidase flux (Cobley & Davison, 2022). The reduction in DSB following both acute and three-week quercetin supplementation suggests more rapid DSB resolution, potentially mediated by SIRT6-dependent chromatin remodelling, PARP1 recruitment, and repair complex assembly at sites of damage (Mao et al., 2011; You et al., 2019). The divergence between DSB foci and comet assay outcomes—where comet measures decreased only after three-week supplementation—likely reflects methodological sensitivity, as super-resolution microscopy detects chromatin-associated foci that conventional comet assays may underestimate. Quercetin may attenuate exercise-induced oxidative DNA damage through both direct antioxidant activity, including radical scavenging and metal chelation that limits Fenton-type reactions, and indirect signalling-dependent mechanisms involving Nrf2 activation and modulation of DNA repair regulators such as PARP1 and SIRT6 (Chen et al., 2021; Yousefi et al., 2024; You et al., 2019).

*SIRT1* expression increased significantly post-exercise in all placebo sessions, consistent with inducible *SIRT1* transcription in response to high-intensity exercise in humans (Juan et al., 2023). This post-exercise induction was absent in the quercetin condition, suggesting attenuation of stress-responsive SIRT1 transcription rather than suppression of SIRT1 function. As a NAD⁺-dependent regulator sensitive to redox and energetic state, SIRT1 transcriptional induction commonly accompanies elevated oxidative or metabolic stress. Quercetin has been shown to enhance SIRT1 enzymatic activity and activate antioxidant pathways across experimental models (Ho et al., 2022; Lu et al., 2023), which may reduce the requirement for inducible SIRT1 expression by stabilising NAD⁺ availability and limiting stress signalling. This interpretation is supported by human skeletal muscle data demonstrating reduced SIRT1 protein abundance but increased deacetylase activity following HIIT, indicating preserved or enhanced function despite lower expression when cellular energetic and redox states are improved (Gurd et al., 2010). Consistent with this framework, quercetin supplementation was associated with reduced DSB burden, supporting preserved SIRT1-mediated repair competence within a less oxidising intracellular environment. Enhanced nuclear Nrf2 translocation with quercetin further supports strengthened endogenous redox-responsive signalling, although antioxidant enzyme activity was not directly measured.

*SIRT6* expression exhibited a contrasting pattern, decreasing post-exercise in placebo sessions but increasing following three-week quercetin supplementation. This response aligns with prior reports of transient *SIRT6* downregulation following exhaustive exercise in humans, reflecting acute oxidative stress signalling rather than sustained loss of repair competence (Radak et al., 2011). Reduced *SIRT6* transcript abundance or nuclear signal under acute stress does not preclude functional engagement, as SIRT6 is rapidly recruited to DSBs where it stabilises chromatin and supports repair through deacetylation-dependent remodelling (Mostoslavsky et al., 2007; Michishita et al., 2008; Chen et al., 2019). Quercetin binds allosterically to SIRT6 and enhances its deacetylase activity, promoting efficient removal of repair-proximal histone acetylation marks and stabilisation of post-repair chromatin states (Kolady & Wang, 2025). Accordingly, preserved or enhanced SIRT6 expression with quercetin is best interpreted as reflecting a reduced oxidative stress environment in which transient stress-associated suppression is alleviated while enzymatic activity is concurrently potentiated, consistent with the observed reduction in DSB burden.

At first glance, increased *SIRT6* expression alongside reduced PARP1 induction with quercetin may appear contradictory, given that SIRT6 activates PARP1 during DNA damage signalling (Mao et al., 2011). However, PARP1 activity is context dependent: while essential for DNA repair, excessive activation under high oxidative stress can become maladaptive due to high NAD⁺ consumption and downstream metabolic strain. Within this framework, absence of *PARP1* induction with quercetin, but not in placebo, is most parsimoniously interpreted as diminished stress-driven activation resulting from lower lesion burden rather than impaired repair capacity, particularly given preserved SIRT6-mediated repair competence. Enhanced Nrf2 nuclear translocation further supports constrained RONS accumulation under supplementation.

*OGG1*, which excises 8-oxoG lesions during BER, decreased post-exercise in placebo sessions but increased following quercetin treatment. Acute repression of *OGG1* in placebo conditions likely reflects transient oxidative stress-induced transcriptional downregulation, as previously observed following exhaustive exercise in humans (Radak et al., 2011). Quercetin may counteract this effect through Nrf2 activation, which directly binds to and upregulates the OGG1 promoter (Shang et al., 2022). Supporting this mechanism, quercetin increased both Nrf2 and OGG1 expression while reducing AP sites and 8-oxodG in rodent models (Darband et al., 2020), and increased OGG1 expression alongside reduced 8-oxodG in human epithelial cells (Min et al., 2009). Together, these findings support Nrf2-mediated preservation of BER competence under reduced oxidative stress, consistent with increased quercetin availability, enhanced nuclear Nrf2 localisation, and reduced oxidative DNA damage observed here.

RAD51, the central recombinase mediating homologous recombination (HR), catalyses homology search and strand invasion during accurate DSB repair. In placebo sessions, *RAD51, SIRT1,* and *PARP1* increased post-exercise, consistent with activation of an oxidative stress-associated DNA damage response. As NAD⁺-dependent chromatin regulators, SIRT1 and PARP1 support damage signalling and repair factor accessibility, facilitating HR engagement under elevated genotoxic stress (Chen et al., 2019). This coordinated induction was absent with quercetin supplementation, consistent with reduced lesion burden and diminished requirement for HR activation under antioxidant and NAD⁺-preserving conditions. Under acute oxidative stress, excessive PARP1 activation can promote PAR-dependent stress responses that constrain energy availability and repair efficiency (Andrabi et al., 2014). Accordingly, reduced *SIRT6* and *OGG1* expression in placebo is best interpreted as transient stress-associated transcriptional suppression rather than loss of repair engagement, coinciding with reliance on stress-responsive SIRT1/PARP1 pathways. In contrast, quercetin preserved and enhanced *SIRT6* and *OGG1* expression, consistent with maintenance of chromatin-associated BER competence under reduced oxidative stress. As HR is typically engaged under higher lesion complexity and imposes greater temporal and regulatory demands than BER, preservation of BER with quercetin may favour a more efficient and metabolically economical repair response in this context.

Glycosylation of plasma proteins and immunoglobulins integrates chronic systemic inflammation, metabolic state, and biological ageing. Pro-inflammatory glycan features, including increased agalactosylation (G0), asialylation (S0), and bisecting GlcNAc (B), are associated with chronic inflammation, whereas increased galactosylation (G1, G2, Gtotal) and sialylation (Stotal) reflect a younger, anti-inflammatory phenotype (Haslund-Gourley et al., 2023). Although HIIE induces transient inflammatory signalling, including cytokine elevations that can influence B-cell activation and glycosylation pathways (Antunes et al., 2019), the directionality of the plasma glycan changes observed here is consistent with an adaptive rather than pathological response.

The coordinated increase in PBMC *PARP1, SIRT1,* and *RAD51* transcription alongside reduced plasma agalactosylation and increased galactosylation suggests parallel engagement of cell-intrinsic genome maintenance pathways and systemic glycome modulation. PARP1, SIRT1, and RAD51 are central components of the DNA damage response and stress adaptation, while plasma N-glycosylation reflects integrated inflammatory and metabolic signalling derived largely from hepatic and immune glycoprotein production. Plasma glycan traits were quantified as relative abundances (percent area), indicating that the observed shift toward increased galactosylation reflects biological remodeling rather than plasma volume effects. Together, these findings are consistent with an acute hormetic response to exercise, in which transient oxidative stress activates DNA repair pathways and is accompanied by modulation of systemic inflammatory tone. At a mechanistic level, RONS can influence glycosylation efficiency through disruption of Golgi pH, disulfide bond formation, and nucleotide sugar transport (Kellokumpu, 2019; Khoder-Agha & Kietzmann, 2021). The attenuation of acute plasma glycan remodelling with quercetin supplementation is therefore consistent with altered engagement of redox-responsive signalling pathways, as indexed by enhanced Nrf2 nuclear translocation, and likely reflects reduced oxidative signalling demand on glycosylation pathways during post-exercise recovery.

In contrast, only non-significant trends were observed for the IgG-derived GlycanAge index, with a tendency to increase post-exercise in placebo and decrease following three-week supplementation. GlycanAge reflects time-integrated biological ageing rather than acute inflammatory responses and is constrained by the long half-life of IgG and the relative stability of its glycosylation under homeostatic conditions (Trzos et al., 2023). Accordingly, GlycanAge is not expected to respond measurably to a single exercise bout or short-term intervention. The absence of significant GlycanAge changes therefore does not contradict the acute plasma glycan shifts observed, but rather underscores the distinction between rapidly responsive plasma glycosylation and more stable IgG-based ageing metrics. While quercetin may favour restoration of youthful IgG glycan structures with sustained exposure, the 21-day supplementation period was likely insufficient to elicit measurable GlycanAge changes, as reported exercise interventions typically span several months (Šimunić-Briški et al., 2024).

In conclusion, HIIE induces transient oxidative DNA damage and dynamic plasma glycan modulation, in contrast to the chronic oxidative and inflammatory stress burden that characterises biological ageing, thereby supporting HIIE as a physiological model of acute oxidative stress. Quercetin supplementation modulated these responses in a manner consistent with improved redox homeostasis and sustained genome maintenance capacity, attenuating post-exercise DNA damage while preserving key repair factors. In parallel, acute plasma glycan remodelling observed under placebo was attenuated with quercetin, suggesting stabilisation of glycosylation dynamics under redox challenge. Collectively, these findings support a framework in which quercetin acts as a context-dependent modulator of repair efficiency, biasing exercise-induced genome maintenance toward energetically economical and chromatin-stabilising pathways rather than damage accumulation.

## Methods

### Participants

Thirteen (n = 13) male participants, between 30-45 years of age (36 ± 4.4 years), of normal weight (BMI 24.5 ± 3.4), light to moderately active (37.6 ± 4.3 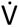O_₂_max) with no medical conditions known to affect responses to exercise and quercetin supplementation took part in this study. Inclusion/exclusion criteria are published elsewhere: https://www.isrctn.com/ISRCTN34136514.

### Quercetin Supplementation

In a randomised, double-blind, placebo-controlled, crossover design, all participants received both quercetin and its placebo counterpart in random order. Each short-term supplementation phase lasted 21 days, during which participants ingested 1000 mg of quercetin aglycone dihydrate (https://DoNotAge.org, UK) or matched placebo capsules once daily with breakfast. Capsules were composed of vegetarian hydroxypropyl methylcellulose (HPMC); the placebo contained rice flour blended with yellow-green food colouring to mimic the appearance of the quercetin capsules. Participants were instructed to refrain from taking additional antioxidant supplements and to maintain their habitual diet and activity patterns throughout the study.

A two to three-week washout period separated the placebo and quercetin treatment phases to ensure complete clearance of quercetin and its metabolites and to minimise carry-over effects. This duration was based on human pharmacokinetic data showing a terminal elimination half-life of approximately 11 h for quercetin glycosides (Graefe et al., 2001) and on bioequivalence guidance recommending that washout intervals exceed about five elimination half-lives to avoid residual effects. Considering quercetin’s enterohepatic recycling and tissue retention, the adopted two to three-week interval corresponds to >30 half-lives, ensuring negligible systemic levels before crossover.

In addition to the short-term supplementation phase, acute supplementation (the first administered dose of quercetin or placebo) was also evaluated. HIIE sessions were conducted at the start (Day 1) and end (Day 21) of each supplementation phase, enabling within-subject comparisons of acute and short-term (21-day) effects of quercetin versus placebo, both before and after exercise-induced oxidative stress. Specifically, the supplement (or its placebo counterpart) was administered 1 hour before exercise, and blood samples were obtained immediately before supplementation and immediately post-exercise. The study design is illustrated in Figure 6.

**Figure 6.**
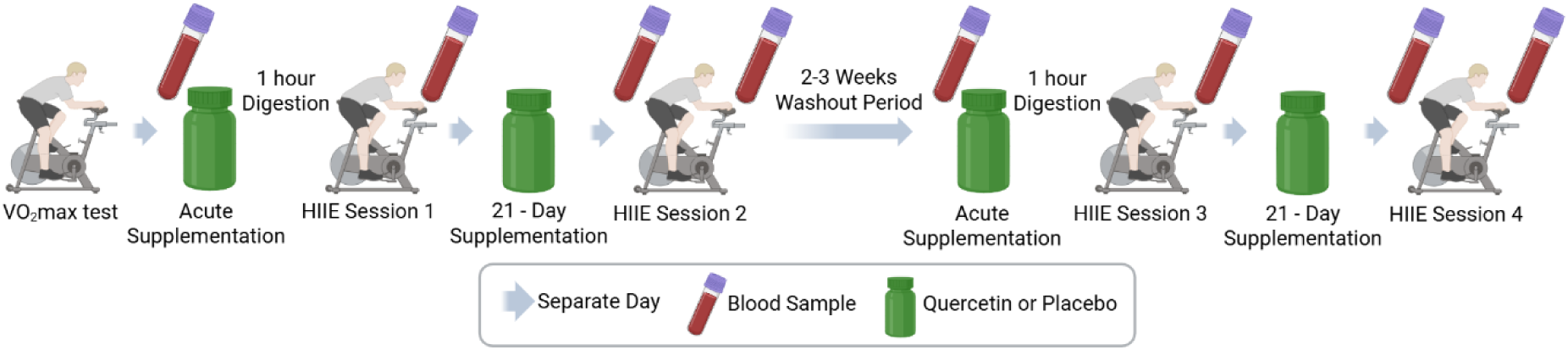
Crossover trial design.

### 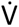O₂max and HIIE

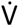O₂max was assessed to prescribe individualised relative exercise intensities for the experimental sessions, ensuring a comparable metabolic load across participants and between time points. Target workloads at 50%, 70%, and 95% of 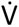O₂max were derived from each participant’s incremental test and used to standardise the warm-up, recovery, and work intervals, respectively. 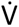O₂max was determined on an electronically braked cycle ergometer (Lode Corival CPET, Lode B.V., Groningen, The Netherlands) using a stepwise ramp protocol in which workload increased by 50 W every 5 min until volitional exhaustion. Breath-by-breath gas exchange was continuously measured (Cosmed Quark CPET, Cosmed, Rome, Italy) following standard two-point gas and flow calibration. 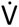O₂ values were time-averaged, and the highest 30-s mean was recorded as 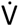O₂max. Individual 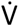O₂ work-rate relationships were plotted in Microsoft Excel and fitted by linear regression to estimate the power outputs corresponding to 50%, 70%, and 95% 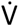O₂max for subsequent exercise sessions. A standardised 4 × 4 min HIIE protocol was then implemented. Each session began with a 10-min warm-up at approximately 50% 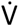O₂max, followed by four 4-min work bouts at approximately 95% 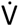O₂max, interspersed with three-minute active recoveries at approximately 70% 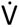O₂max. The prescribed powers for the warm-up, recovery, and work intervals were taken from each participant’s regression-derived targets to ensure intensity precision and reproducibility across both the acute and short-term (day 21) testing phases.

### Haematology

Blood was drawn from the antecubital fossa of the non-dominant arm using a 21-gauge butterfly needle while participants were in a supine position to minimise venous pooling and physiological variability. Samples were collected into a combination of EDTA, serum-separator, and PAXgene RNA vacutainers (BD Biosciences, Franklin Lakes, NJ, USA; PreAnalytiX, Hombrechtikon, Switzerland) to allow downstream molecular and biochemical analyses. EDTA and serum-separator tubes were centrifuged within 30 min of collection at 1500 × g for 10 min at 4 °C. Plasma and serum were aliquoted (1 mL) into sterile cryovials and stored at −80 °C until analysis of oxidative stress and glycomic markers. Whole blood collected in PAXgene tubes was gently inverted 10 times immediately after collection, kept at room temperature for 2 h to ensure complete lysis, then stored at −20 °C overnight before transfer to −80 °C for long-term storage pending RNA extraction. All samples were labelled with anonymised participant IDs, treatment phase, and time point, and were maintained on ice throughout handling. Freeze-thaw cycles were avoided to preserve molecular integrity.

In addition to venous sampling, arterialised capillary blood was obtained by finger-prick for the determination of haemoglobin and haematocrit using the equations described by Dill and Costill (1974) (Supplementary Table S1). All collections were performed between 09:00 and 10:00 to minimise diurnal variation in molecular responses to exercise. Immediately following baseline blood sampling, participants commenced the HIIE session. Post-exercise blood draws were collected immediately after cooldown.

### Plasma Quercetin, Cell Uptake, and Colocalisation with SIRT6

Blood quercetin aglycone concentrations were determined using ultra-high-performance liquid chromatography coupled with tandem mass spectrometry (UHPLC–MS/MS), as previously described for plasma flavonol quantification in human supplementation studies. A total of 100 µL plasma was mixed with 300 µL acetonitrile (≥99.9 %, LC–MS grade; Sigma-Aldrich, St Louis, MO, USA) to precipitate proteins, vortex-mixed for 30 s, and centrifuged for 15 min at 1500 × g, 4 °C. The supernatant was extracted with 300 µL ethyl acetate (≥99.8 %, Sigma-Aldrich), vortex-mixed for 30 s, and centrifuged again under identical conditions. 250 µL of the resulting supernatant was transferred to a 5 mL borosilicate glass vial (Agilent Technologies, Waldbronn, Germany) and evaporated to dryness under a gentle stream of nitrogen using a Techne Dri-Block DB-3D sample concentrator (Techne, Staffordshire, UK) at room temperature for 30 min. The residue was reconstituted in 100 µL acetonitrile, sonicated for 10 s, vortex-mixed for 30 s, and transferred to UHPLC vials for analysis. All samples and standards were processed in parallel under subdued light to minimise quercetin degradation.

Samples were analysed using an Agilent 1200 Series HPLC system (Agilent Technologies, Santa Clara, CA, USA) coupled to a Thermo Scientific TSQ Vantage triple quadrupole mass spectrometer (Thermo Fisher Scientific, San Jose, CA, USA), with a negative electrospray ionisation (ESI) mode using multiple reaction monitoring (MRM). Chromatographic separation was achieved on a ZORBAX Eclipse XDB-C18 column (1.8 µm, 4.6 x 50 mm; Agilent Technologies, Santa Clara, CA, USA) maintained at 30 °C. The mobile phases consisted of (A) water with 0.1 % (v/v) formic acid (Honeywell, Charlotte, NC, USA) and (B) acetonitrile (LC–MS grade; Sigma-Aldrich). Typical MS source parameters were: spray voltage −4500 V, sheath gas 50 (arbitrary units), auxiliary gas 60 (arbitrary units), and source temperature 350 °C. MRM transitions were monitored for quercetin aglycone (precursor m/z 301.0 → product m/z 151.0 and 179.0; dwell time 50 ms per transition). Peak areas were integrated using Xcalibur (version 4.3; Thermo Fisher Scientific, San Jose, CA, USA) and calibration curves were generated by weighted (1/x) linear regression in Microsoft Excel.

To assess intracellular uptake of native quercetin and its colocalisation with SIRT6, PBMCs collected before and after quercetin and placebo supplementation were analysed using super-resolution 3D STED confocal microscopy, following established protocols for quercetin autofluorescence and SIRT6 immunofluorescence (Drummond et al., 2017; Pan et al., 2016). Cells were stored at −80 °C in cryopreservation medium containing 10 % dimethyl sulfoxide (DMSO; Sigma-Aldrich, St Louis, MO, USA) and fetal bovine serum (FBS; Gibco, Thermo Fisher Scientific, Waltham, MA, USA). For analysis, cells were thawed in a 37 °C water bath, diluted in pre-warmed RPMI-1640 medium (Gibco) containing 10 % FBS, and centrifuged to remove DMSO. A total of 200 µL lymphocytes was spotted onto Superfrost Plus slides (Thermo Fisher Scientific) and allowed to adhere for 15 min. Cells were fixed with 4 % paraformaldehyde in PBS (Sigma-Aldrich) for 10 min, washed three times in PBS (Sigma-Aldrich), and permeabilised with 0.5 % Triton X-100/PBS (Sigma-Aldrich) for 10 min. Non-specific binding was blocked with 2.5 % normal horse serum (NHS; Vector Laboratories, Burlingame, CA, USA) for 1 h. Slides were then incubated with rabbit polyclonal anti-SIRT6 antibody (1:500; Proteintech, 28677-1-AP, Manchester, UK) in 1 % NHS/PBS for 1 h at room temperature, washed three times in 0.05 % Tween-20/1 % NHS/PBS, and incubated with goat anti-rabbit Alexa Fluor 568 antibody (1:500; Invitrogen, Thermo Fisher Scientific) for 1 h.

Quercetin autofluorescence was visualised directly in the green channel (excitation 488 nm), eliminating the need for staining. Nuclei were counterstained with Vectashield Antifade Mounting Medium containing DAPI (Vector Laboratories) and coverslipped with clear nail polish. 3D images were acquired using a Leica Stellaris One Tau 3D STED Nanoscope (Leica Microsystems, Wetzlar, Germany) with a 100×/1.4 NA oil-immersion objective. Excitation/emission settings were: DAPI (Ex 405 nm, Em 415–480 nm; gain 5 %, laser 10 %), Alexa Fluor 488 (Ex 488 nm, Em 500–550 nm; gain 30 %, laser 100 %), and Alexa Fluor 568 (Ex 561 nm, Em 570–620 nm; gain 30 %, laser 70 %). Sequential scanning was used to prevent spectral cross-talk. For each field of view, Z-stacks were captured spanning the full nuclear depth, with all imaging parameters (laser power, detector gain, and pinhole size) held constant. Each image contained ≥ 40 nuclei, and at least three independent fields (≥ 100 nuclei total) were analysed per condition. 3D image stacks were processed using Imaris 10.2 (Oxford Instruments, UK). Raw STED data were converted to Imaris format and subjected to automated colocalisation analysis.

For colocalisation analysis with SIRT6, thresholds for quercetin (green; Alexa Fluor 488) and SIRT6 (red; Alexa Fluor 568) channels were set to 90, and for DAPI to 20, based on the faintest visible foci within each batch. Nuclear volumes were segmented from the DAPI channel using the Surface module, and spot detection was applied separately to quercetin and SIRT6 channels. SIRT6-quercetin colocalisation was quantified using the Colocalisation module, reporting the percentage region of interest (%ROI)—the proportion of the nuclear volume containing overlapping quercetin and SIRT6 signals—as the primary outcome measure. This voxel-based 3D quantification approach was adapted from established Imaris colocalisation workflows (Pradhan et al., 2022). Median fluorescence intensities for green and red channels were quantified for each nucleus using automated batch processing tools in Imaris. All imaging and analysis parameters (laser power, detector gain, thresholds, segmentation) were held constant across experimental conditions to ensure comparability. Secondary-only and no-primary antibody controls confirmed antibody specificity.

### DNA Damage and Nrf2 Localisation

DNA strand breaks were quantified using the alkaline comet assay, performed according to the protocol of Collins et al. (2023). Oxidised purine lesions were assessed using a modified version of this assay incorporating formamidopyrimidine-DNA glycosylase (FPG). In brief, the standard alkaline comet assay procedure was followed, with the additional step of FPG incubation to convert oxidised purines into detectable strand breaks, as described by Collins et al. (2023).

For DNA DSB and Nrf2 localisation, immunofluorescence with super-resolution 3D microscopy was performed. DSBs were visualised through γH2AX and 53BP1 co-localisation, while Nrf2 nuclear localisation was assessed by median intensity. Super-resolution 3D confocal and STED imaging provides superior sensitivity for detecting γH2AX/53BP1 foci compared with 2D microscopy, as 2D projections can underestimate DSB foci by ∼40 % (Jezkova et al., 2018) and confocal microscopy with 3D reconstruction identifies approximately threefold more foci (Ruprecht et al., 29). A total of 200 µL lymphocytes was spotted onto Superfrost Plus slides (Fisherbrand, Thermo Fisher Scientific, USA) and allowed to adhere for 15 min. Cells were fixed with 4% paraformaldehyde in PBS (Sigma-Aldrich, USA) for 10 min, washed three times in PBS, and permeabilised with 0.5 % Triton X-100/PBS (Sigma-Aldrich) for 10 min. Slides were blocked with 2.5 % normal horse serum (NHS; Vector Laboratories, USA) for 1 h, then incubated for 1 h with either: anti-phospho-histone H2AX (Ser139, mouse mAb, 1:500; Invitrogen, Thermo Fisher Scientific) together with anti-53BP1 (rabbit pAb, 1:500; Invitrogen, Thermo Fisher Scientific) for DSB analysis, or CoraLite® 594-conjugated anti-Nrf2 (rabbit pAb, 1:500; Proteintech 16396-1-AP, Manchester, UK) for Nrf2 localisation, following established protocols for oxidative stress-induced Nrf2 activation. Slides were washed three times in 0.05 % Tween-20/1 % NHS/PBS (Sigma-Aldrich).

For DSB imaging, cells were further incubated with goat anti-mouse Alexa Fluor® 488 IgG (1:500; Invitrogen A-11001) and goat anti-rabbit Alexa Fluor® 568 IgG (1:500; Invitrogen A-11011) for 1 h. Slides were mounted with Vectashield Antifade with DAPI (Vector Laboratories) and sealed with clear nail varnish. Images were acquired using a Leica Stellaris One Tau 3D STED Nanoscope (Leica Microsystems, Germany) with a 100×/1.4 NA oil-immersion objective. Sequential scanning was used to prevent spectral overlap using the following settings for DSB analysis: DAPI (excitation 405 nm; emission 415–480 nm; detector gain 5%, laser power 10%), Alexa Fluor 488 (excitation 488 nm; emission 500–550 nm; detector gain 30%, laser power 70%), and Alexa Fluor 568 (excitation 561 nm; emission 570–620 nm; detector gain 30%, laser power 70%). For Nrf2: DAPI (excitation 405 nm; emission 415–480 nm; detector gain 5%, laser power 10%) and Alexa Fluor 594 (excitation 593 nm; emission 610–620 nm; detector gain 30%, laser power 70%). Laser power, detector gain, and pinhole size were kept constant. Each field contained ≥ 40 nuclei, with ≥ 3 fields per sample (≥ 100 nuclei total).

Z-stacks were processed in Imaris 10.2 (Oxford Instruments, UK). For DSB analysis, thresholds were set to 1000 (green/red) and 50 (DAPI). Nuclear volumes were segmented from DAPI, and colocalisation was quantified as %ROI. For Nrf2, median intensity per nucleus was determined by automated batch analysis. All imaging and analysis parameters were held constant across conditions. Secondary-only and no-primary antibody controls confirmed antibody specificity.

### DNA Repair

Whole blood was collected in PAXgene® Blood RNA Tubes (PreAnalytiX GmbH, Hombrechtikon, Switzerland; distributed by Qiagen, Hilden, Germany) to stabilise intracellular RNA immediately after phlebotomy. Total RNA was isolated using the PAXgene® Blood RNA Kit (Qiagen, Hilden, Germany) according to the manufacturer’s instructions, including on-column DNase digestion to remove residual genomic DNA. RNA concentration and purity were assessed spectrophotometrically using a NanoDrop™ ND-1000 Spectrophotometer (Thermo Fisher Scientific, Waltham, MA, USA) by measuring A260/A280 ratios.

RNA samples were diluted with nuclease-free water (Ambion, Thermo Fisher Scientific) to ensure exactly 500 ng of total RNA (or up to 1 µg where indicated) per reverse-transcription reaction. Complementary DNA (cDNA) was synthesised using the High-Capacity cDNA Reverse Transcription Kit (Applied Biosystems, Foster City, CA, USA) in a 20 µL reaction (25 °C for 10 min, 37 °C for 120 min, 85 °C for 5 min) following the manufacturer’s protocol. Synthesised cDNA was stored at –20 °C until further use. Each 20 µL qPCR reaction contained 10 µL TaqMan® Universal PCR Master Mix (2×) (Applied Biosystems), 1 µL cDNA template (≈ 25 ng reverse-transcribed RNA), 0.5 µL forward primer (10 µM; final ≈ 250 nM), 0.5 µL reverse primer (10 µM; ≈ 250 nM), 0.5 µL dual-labelled hydrolysis probe (5 µM), and 7.5 µL nuclease-free water (Ambion, Thermo Fisher Scientific). Amplifications were carried out in 96-well optical plates on an Applied Biosystems 7500 Fast Real-Time PCR System using standard cycling conditions: 95 °C for 10 min, followed by 40 cycles of 95 °C for 15 s and 60 °C for 1 min.

All reactions were performed in technical duplicates, and no-template controls were included to verify the absence of contamination. Gene expression was assessed for six DNA repair-related targets using TaqMan® Gene Expression Assays (Applied Biosystems), with half of the probes labelled with VIC® dye and half with FAM™ dye for multiplex detection. Expression data were normalised to the geometric mean of three housekeeping genes (*HPRT1, TUBB, GAPDH*) to account for differences in RNA input and amplification efficiency.Primers and probes (5′→3′):

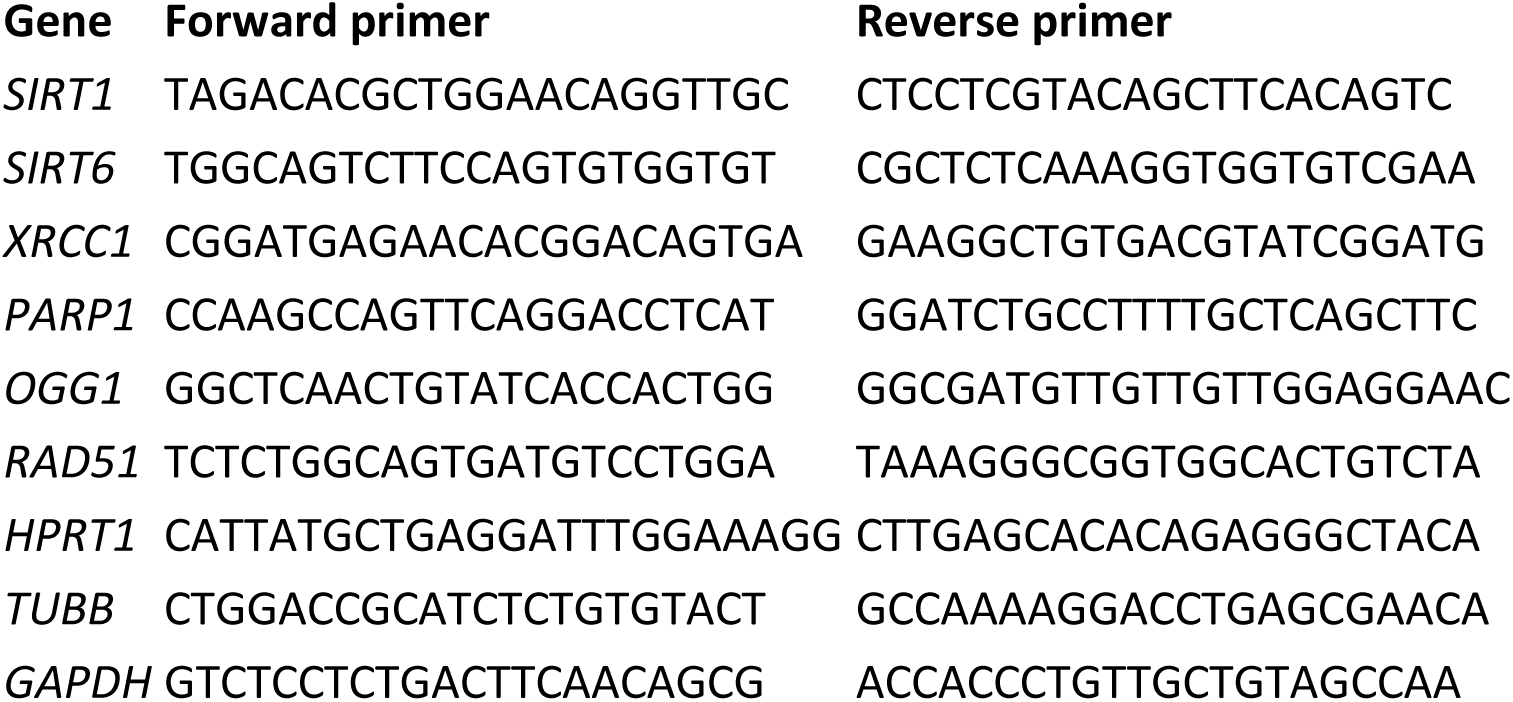

Relative gene expression levels were quantified using the ΔCt method, with target gene expression normalised to the geometric mean of three reference genes (HPRT, TUBB, and GAPDH). ΔCt values were exported to IBM SPSS Statistics (Version 31; IBM Corp., Armonk, NY, USA) for statistical analyses. Fold changes in gene expression were calculated using the 2^(−ΔΔCt)^ method and visualised as bar graphs.

### GlycanAge, IgG and plasma N-glycan profiling

IgG N-glycans were analysed using a capillary gel electrophoresis with laser-induced fluorescence (CGE-LIF) developed by Hanić et al. (2019). The analysis was performed by isolating IgG from plasma samples, using a Protein G monolithic plate in detail described in the protocol, followed by the release and fluorescent labelling of N-glycans. N-glycans were incubated with PNGase F and released from denatured glycoproteins, labelled with 8-aminopyrene-1,3,6-trisulfonic acid trisodium salt (APTS) and purified from salt and excess label by HILIC-based solid phase extraction (HILIC-SPE) (Ruhaak et al., 2010). CGE-LIF glycoprofiling was performed using a 3130 Genetic Analyser (Applied Biosystems, Waltham, MA, USA). The obtained electropherograms were integrated into 27 peaks using Waters Empower 3 software (Waters, US). The list of the corresponding structures for these peaks is provided in the Supplementary Table S2.

Plasma glycans were isolated from individual plasma samples, denatured by incubation with SDS (Invitrogen, Waltham, MA, USA) at 65 °C for 10 min, followed by the release of glycans with the addition of 1.2 units of peptide: N-glycosidase F (Promega, Madison, MI, USA) and overnight incubation at 37 °C. N-glycans were labelled with 2-aminobenzamide (Sigma-Aldrich, Saint Louis, MO, USA), separated by hydrophilic interaction liquid chromatography on an Acquity Ultra-Performance Liquid Chromatography (UPLC Technology) instrument (Waters, Milford, MA, US), and quantified with a fluorescence detector using the hydrophilic interaction ultra-high-performance liquid chromatography with fluorescence detection (HILIC-UHPLC-FLR) method. Chromatograms were separated into 39 glycan peaks (GPs) (Supplementary Table S2).

GlycanAge is an index of biological age that estimates how old the body is functionally by analysing the N-glycans attached to IgG—the main antibody circulating in plasma. These IgG glycans modulate antibody effector functions and systemic inflammation, and their composition changes predictably with age. GlycanAge measures inflammageing, the chronic, low-grade inflammation that develops progressively with age, even in the absence of infection, and represents a central integrative hallmark that drives or amplifies many other hallmarks of ageing. GlycanAge was derived from IgG glycan peaks quantified using the protocol described above.

### Statistical Analysis

Data normality was assessed using the Shapiro-Wilk test, and non-normal data were log-transformed prior to analysis; when normality was not achieved, non-parametric tests were applied. Primary analyses used a repeated-measures general linear model with Time (pre vs post-exercise) and Treatment condition (quercetin vs placebo) to test Time × Treatment interactions. Significant interactions (p < 0.05) were explored using post-hoc one-way ANOVA with Tukey correction (or Kruskal–Wallis tests for non-parametric data) to compare between-condition differences at individual timepoints. Within-condition pre vs post-exercise comparisons were performed using paired t-tests (or Wilcoxon signed-rank tests for non-parametric data). To control the family-wise error rate, a Bonferroni correction with m = 2 was applied within each treatment period, corresponding to the two pre-post exercise contrasts (Day 1 and Day 21). Because treatment periods were separated by a sufficient washout to minimise carry-over, multiplicity was controlled within periods rather than across all four contrasts. Results are reported as mean ± SD for normally distributed data and median (IQR) for non-normal data.

### Ethical Approval

The human trial was approved by Ulster University Research Ethics Committee (REC/23/0020) and registered in a publicly accessible database recognised by the World Health Organization (ISRCTN34136514).

## Acknowledgements

The authors thank Genos Ltd. (Zagreb, Croatia) for performing the glycomic profiling of IgG and plasma proteins.

## Author Contributions

Investigation & Formal Analysis: C.G.J.; Formal Analysis & Resources: N.S.B., F.V.Z.; Supervision: G.D., K.M.

## Data Availability Statement

The data that support the findings of this study are available from the corresponding author upon reasonable request.

## Supplementary Materials

**Supplementary Table S1.**
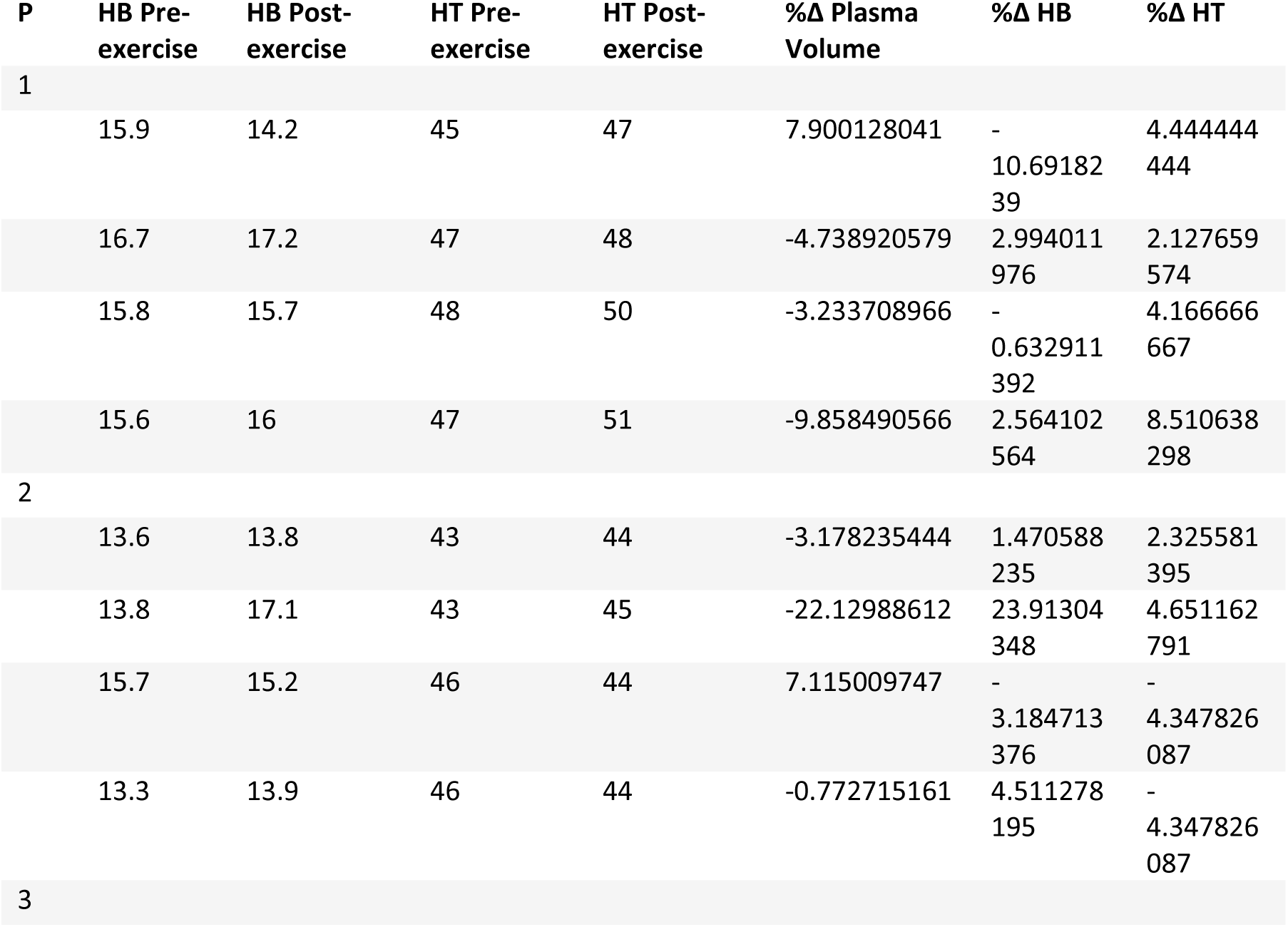
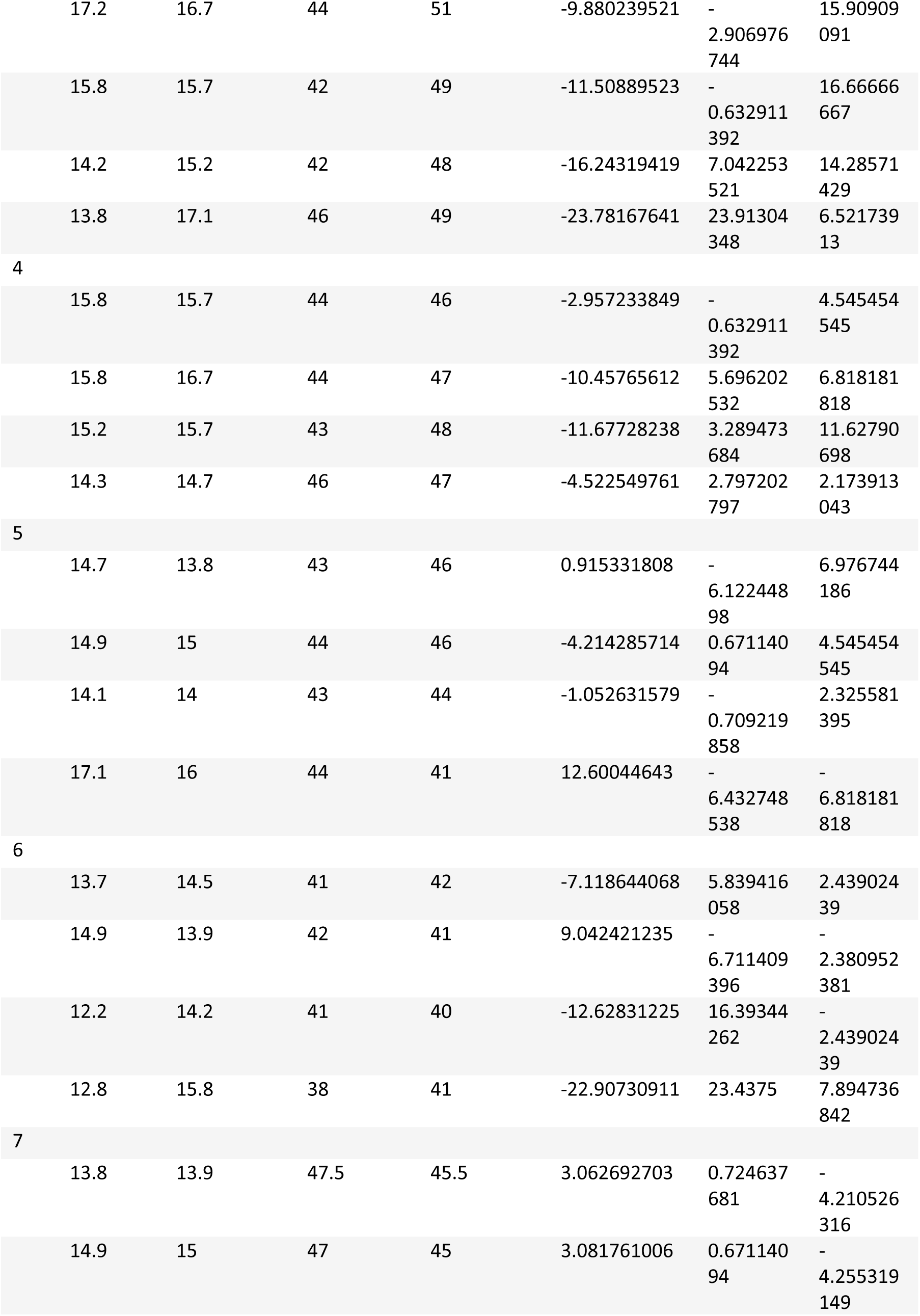
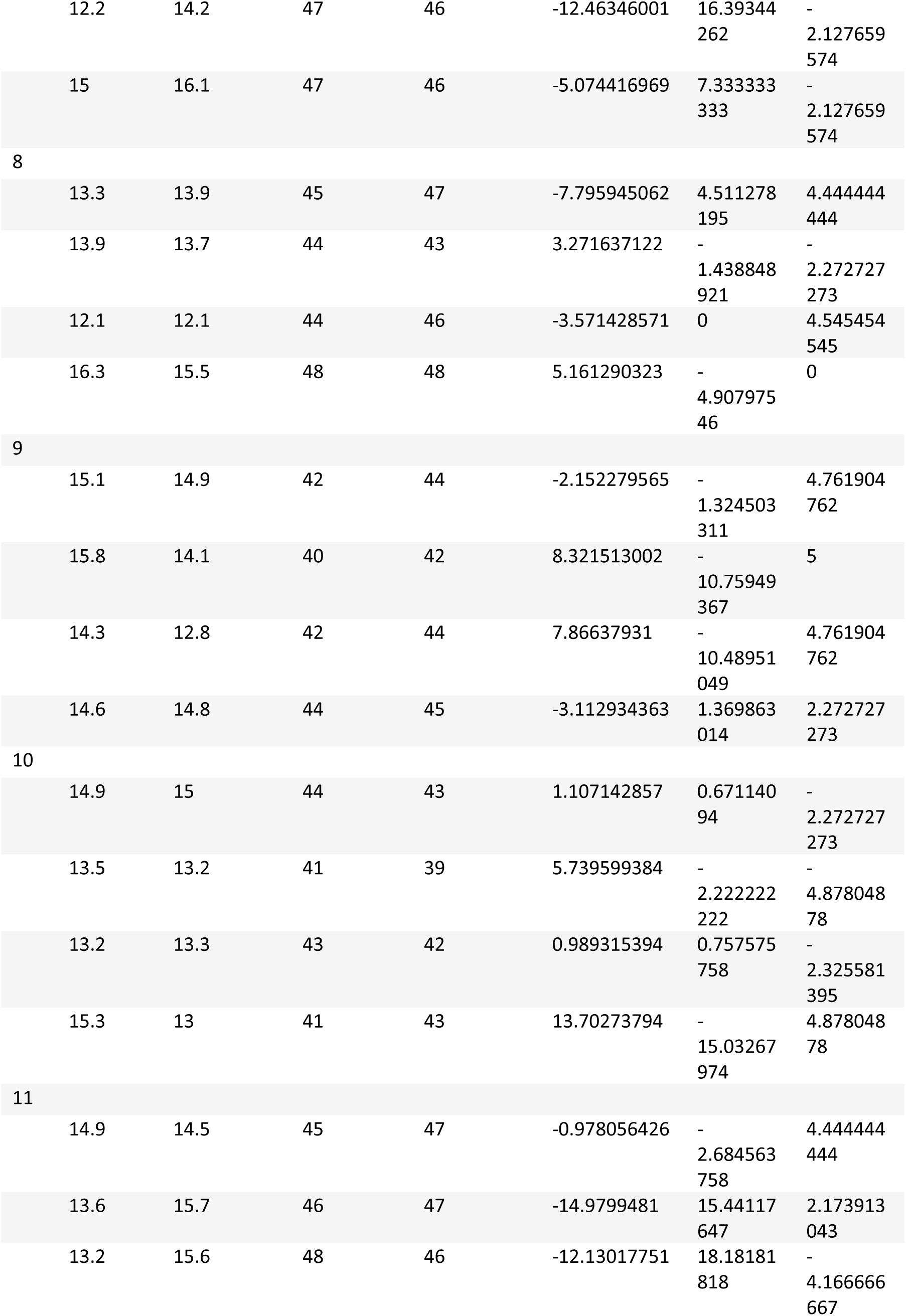
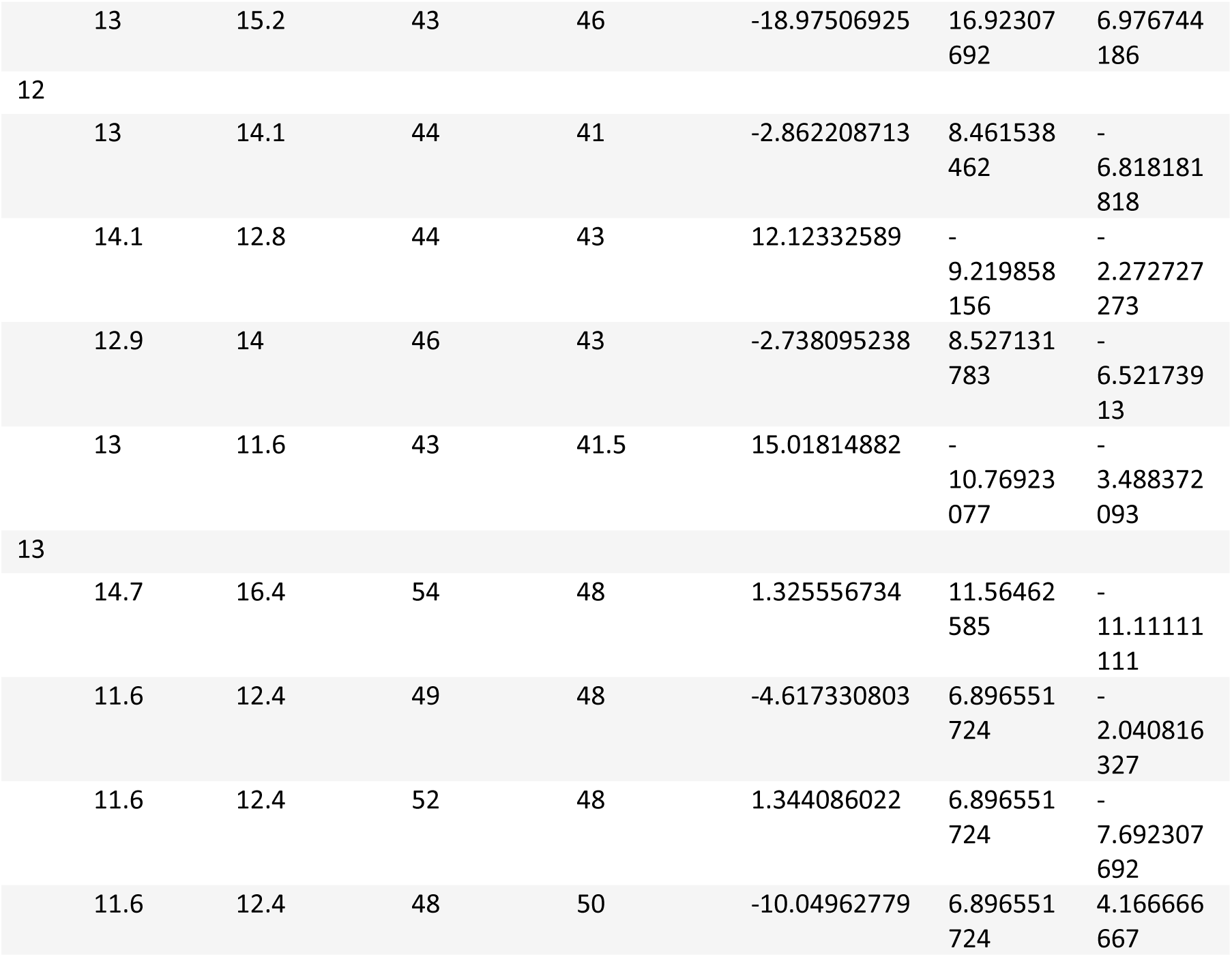
Haemoglobin, haematocrit, and estimated plasma volume changes before and after acute exercise. HB haemoglobin; HT haematocrit; P participant.

| P | HB Pre-exercise | HB Post-exercise | HT Pre-exercise | HT Post-exercise | %Δ Plasma Volume | %Δ HB | %Δ HT |
| --- | --- | --- | --- | --- | --- | --- | --- |
| 1 |  |  |  |  |  |  |  |
|  | 15.9 | 14.2 | 45 | 47 | 7.900128041 | -<br>10.69182<br>39 | 4.444444<br>444 |
|  | 16.7 | 17.2 | 47 | 48 | -4.738920579 | 2.994011<br>976 | 2.127659<br>574 |
|  | 15.8 | 15.7 | 48 | 50 | -3.233708966 | -<br>0.632911<br>392 | 4.166666<br>667 |
|  | 15.6 | 16 | 47 | 51 | -9.858490566 | 2.564102<br>564 | 8.510638<br>298 |
| 2 |  |  |  |  |  |  |  |
|  | 13.6 | 13.8 | 43 | 44 | -3.178235444 | 1.470588<br>235 | 2.325581<br>395 |
|  | 13.8 | 17.1 | 43 | 45 | -22.12988612 | 23.91304<br>348 | 4.651162<br>791 |
|  | 15.7 | 15.2 | 46 | 44 | 7.115009747 | -<br>3.184713<br>376 | -<br>4.347826<br>087 |
|  | 13.3 | 13.9 | 46 | 44 | -0.772715161 | 4.511278<br>195 | -<br>4.347826<br>087 |
| 3 |  |  |  |  |  |  |  |

|  |  |  |  |  |  |  |
| --- | --- | --- | --- | --- | --- | --- |
| 17.2 | 16.7 | 44 | 51 | -9.880239521 | -<br>2.906976<br>744 | 15.90909<br>091 |
| 15.8 | 15.7 | 42 | 49 | -11.50889523 | -<br>0.632911<br>392 | 16.66666<br>667 |
| 14.2 | 15.2 | 42 | 48 | -16.24319419 | 7.042253<br>521 | 14.28571<br>429 |
| 13.8 | 17.1 | 46 | 49 | -23.78167641 | 23.91304<br>348 | 6.521739<br>13 |
| 4 |  |  |  |  |  |  |
| 15.8 | 15.7 | 44 | 46 | -2.957233849 | -<br>0.632911<br>392 | 4.545454<br>545 |
| 15.8 | 16.7 | 44 | 47 | -10.45765612 | 5.696202<br>532 | 6.818181<br>818 |
| 15.2 | 15.7 | 43 | 48 | -11.67728238 | 3.289473<br>684 | 11.62790<br>698 |
| 14.3 | 14.7 | 46 | 47 | -4.522549761 | 2.797202<br>797 | 2.173913<br>043 |
| 5 |  |  |  |  |  |  |
| 14.7 | 13.8 | 43 | 46 | 0.915331808 | -<br>6.122448<br>98 | 6.976744<br>186 |
| 14.9 | 15 | 44 | 46 | -4.214285714 | 0.671140<br>94 | 4.545454<br>545 |
| 14.1 | 14 | 43 | 44 | -1.052631579 | -<br>0.709219<br>858 | 2.325581<br>395 |
| 17.1 | 16 | 44 | 41 | 12.60044643 | -<br>6.432748<br>538 | -<br>6.818181<br>818 |
| 6 |  |  |  |  |  |  |
| 13.7 | 14.5 | 41 | 42 | -7.118644068 | 5.839416<br>058 | 2.439024<br>39 |
| 14.9 | 13.9 | 42 | 41 | 9.042421235 | -<br>6.711409<br>396 | -<br>2.380952<br>381 |
| 12.2 | 14.2 | 41 | 40 | -12.62831225 | 16.39344<br>262 | -<br>2.439024<br>39 |
| 12.8 | 15.8 | 38 | 41 | -22.90730911 | 23.4375 | 7.894736<br>842 |
| 7 |  |  |  |  |  |  |
| 13.8 | 13.9 | 47.5 | 45.5 | 3.062692703 | 0.724637<br>681 | -<br>4.210526<br>316 |
| 14.9 | 15 | 47 | 45 | 3.081761006 | 0.671140<br>94 | -<br>4.255319<br>149 |
| 12.2 | 14.2 | 47 | 46 | -12.46346001 | 16.39344<br>262 | -<br>2.127659<br>574 |
| 15 | 16.1 | 47 | 46 | -5.074416969 | 7.333333<br>333 | -<br>2.127659<br>574 |
| 8 |  |  |  |  |  |  |
| 13.3 | 13.9 | 45 | 47 | -7.795945062 | 4.511278<br>195 | 4.444444<br>444 |
| 13.9 | 13.7 | 44 | 43 | 3.271637122 | -<br>1.438848<br>921 | -<br>2.272727<br>273 |
| 12.1 | 12.1 | 44 | 46 | -3.571428571 | 0 | 4.545454<br>545 |
| 16.3 | 15.5 | 48 | 48 | 5.161290323 | -<br>4.907975<br>46 | 0 |
| 9 |  |  |  |  |  |  |
| 15.1 | 14.9 | 42 | 44 | -2.152279565 | -<br>1.324503<br>311 | 4.761904<br>762 |
| 15.8 | 14.1 | 40 | 42 | 8.321513002 | -<br>10.75949<br>367 | 5 |
| 14.3 | 12.8 | 42 | 44 | 7.86637931 | -<br>10.48951<br>049 | 4.761904<br>762 |
| 14.6 | 14.8 | 44 | 45 | -3.112934363 | 1.369863<br>014 | 2.272727<br>273 |
| 10 |  |  |  |  |  |  |
| 14.9 | 15 | 44 | 43 | 1.107142857 | 0.671140<br>94 | -<br>2.272727<br>273 |
| 13.5 | 13.2 | 41 | 39 | 5.739599384 | -<br>2.222222<br>222 | -<br>4.878048<br>78 |
| 13.2 | 13.3 | 43 | 42 | 0.989315394 | 0.757575<br>758 | -<br>2.325581<br>395 |
| 15.3 | 13 | 41 | 43 | 13.70273794 | -<br>15.03267<br>974 | 4.878048<br>78 |
| 11 |  |  |  |  |  |  |
| 14.9 | 14.5 | 45 | 47 | -0.978056426 | -<br>2.684563<br>758 | 4.444444<br>444 |
| 13.6 | 15.7 | 46 | 47 | -14.9799481 | 15.44117<br>647 | 2.173913<br>043 |
| 13.2 | 15.6 | 48 | 46 | -12.13017751 | 18.18181<br>818 | -<br>4.166666<br>667 |
| 13 | 15.2 | 43 | 46 | -18.97506925 | 16.92307<br>692 | 6.976744<br>186 |
| 12 |  |  |  |  |  |  |
| 13 | 14.1 | 44 | 41 | -2.862208713 | 8.461538<br>462 | -<br>6.818181<br>818 |
| 14.1 | 12.8 | 44 | 43 | 12.12332589 | -<br>9.219858<br>156 | -<br>2.272727<br>273 |
| 12.9 | 14 | 46 | 43 | -2.738095238 | 8.527131<br>783 | -<br>6.521739<br>13 |
| 13 | 11.6 | 43 | 41.5 | 15.01814882 | -<br>10.76923<br>077 | -<br>3.488372<br>093 |
| 13 |  |  |  |  |  |  |
| 14.7 | 16.4 | 54 | 48 | 1.325556734 | 11.56462<br>585 | -<br>11.11111<br>111 |
| 11.6 | 12.4 | 49 | 48 | -4.617330803 | 6.896551<br>724 | -<br>2.040816<br>327 |
| 11.6 | 12.4 | 52 | 48 | 1.344086022 | 6.896551<br>724 | -<br>7.692307<br>692 |
| 11.6 | 12.4 | 48 | 50 | -10.04962779 | 6.896551<br>724 | 4.166666<br>667 |

**Supplementary Table S2A.**
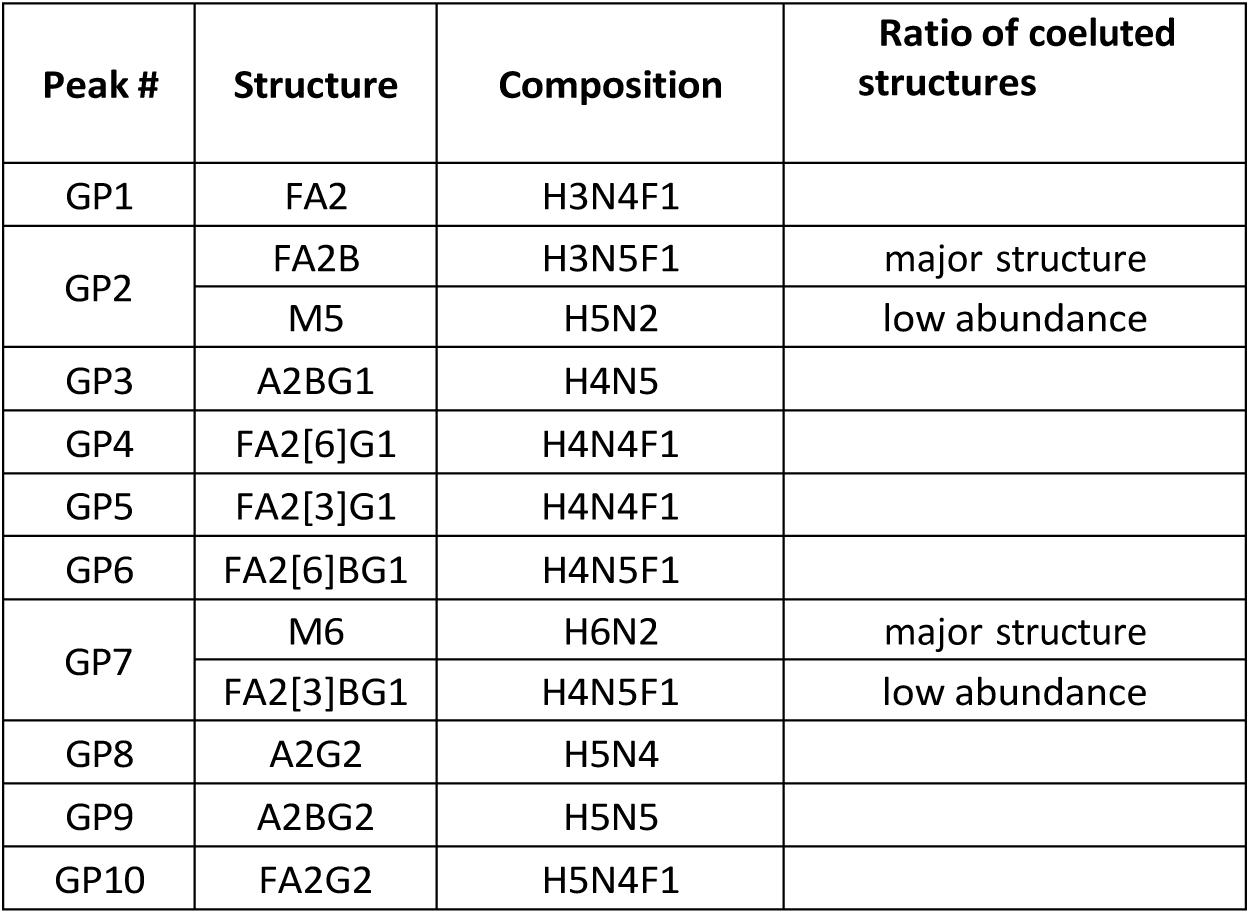
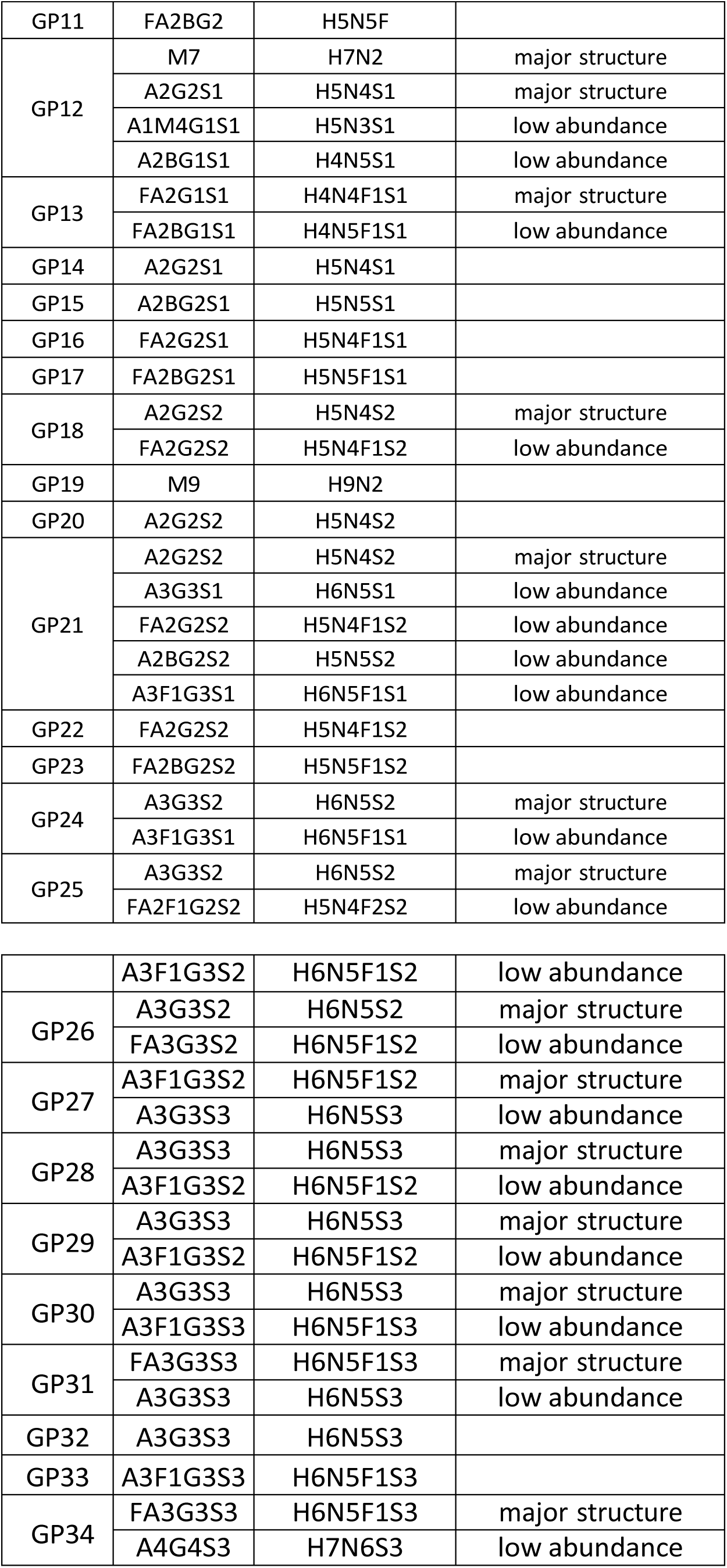
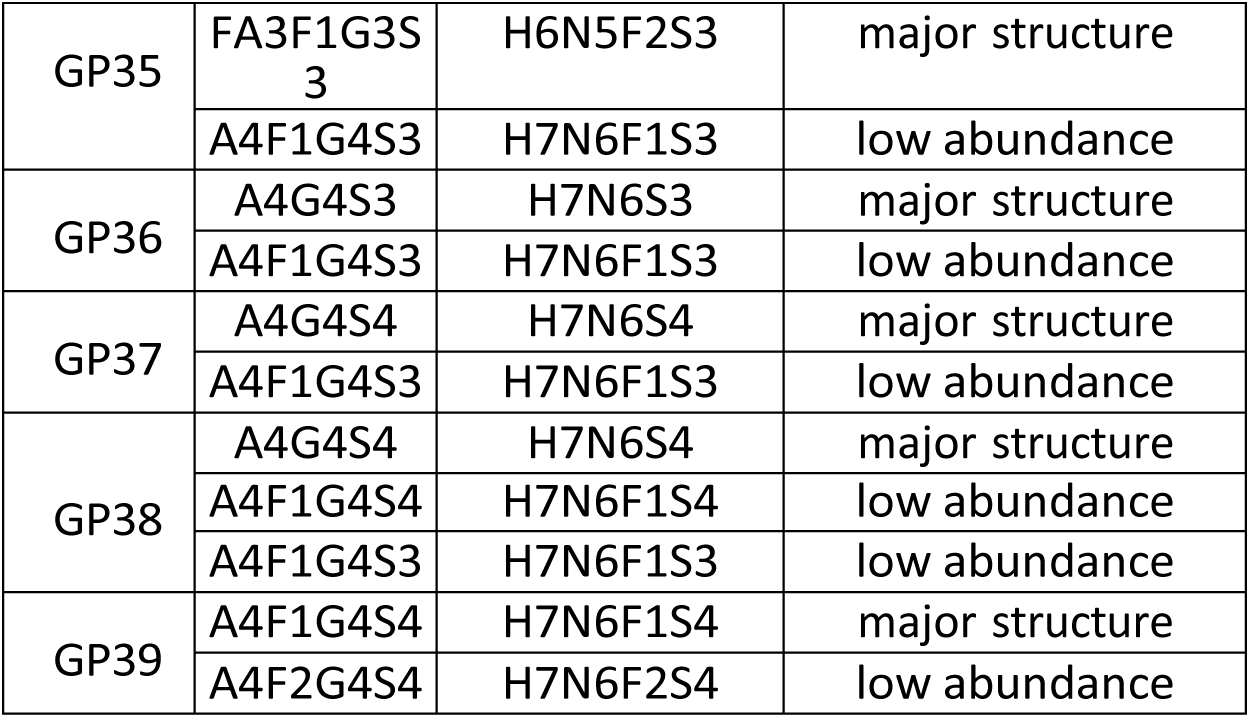
The glycan structures corresponding to each of the glycan peaks (GP) are shown. In the case of multiple structures per glycan peak, the upper structure is the major one, and the lower one is minor in abundance.

**Supplementary Table S2B.**
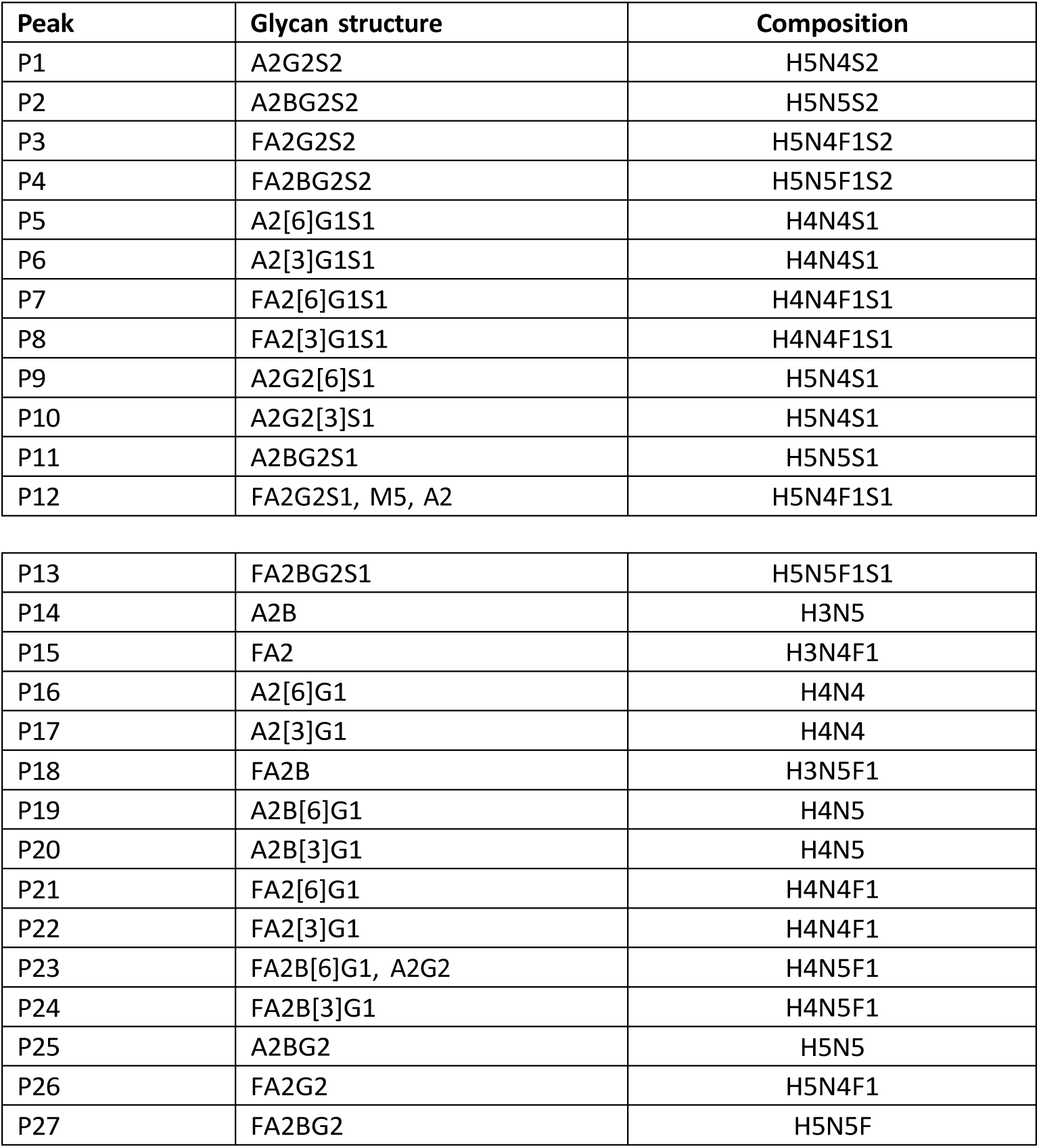
The glycan structures corresponding to each of the IgG glycan peaks in CGE (P) are shown.

| Peak | Glycan structure | Composition |
| --- | --- | --- |
| P1 | A2G2S2 | H5N4S2 |
| P2 | A2BG2S2 | H5N5S2 |
| P3 | FA2G2S2 | H5N4F1S2 |
| P4 | FA2BG2S2 | H5N5F1S2 |
| P5 | A2[6]G1S1 | H4N4S1 |
| P6 | A2[3]G1S1 | H4N4S1 |
| P7 | FA2[6]G1S1 | H4N4F1S1 |
| P8 | FA2[3]G1S1 | H4N4F1S1 |
| P9 | A2G2[6]S1 | H5N4S1 |
| P10 | A2G2[3]S1 | H5N4S1 |
| P11 | A2BG2S1 | H5N5S1 |
| P12 | FA2G2S1, M5, A2 | H5N4F1S1 |
| P13 | FA2BG2S1 | H5N5F1S1 |
| P14 | A2B | H3N5 |
| P15 | FA2 | H3N4F1 |
| P16 | A2[6]G1 | H4N4 |
| P17 | A2[3]G1 | H4N4 |
| P18 | FA2B | H3N5F1 |
| P19 | A2B[6]G1 | H4N5 |
| P20 | A2B[3]G1 | H4N5 |
| P21 | FA2[6]G1 | H4N4F1 |
| P22 | FA2[3]G1 | H4N4F1 |
| P23 | FA2B[6]G1, A2G2 | H4N5F1 |
| P24 | FA2B[3]G1 | H4N5F1 |
| P25 | A2BG2 | H5N5 |
| P26 | FA2G2 | H5N4F1 |
| P27 | FA2BG2 | H5N5F |

**Supplementary Table S3.** Plasma glycan changes from pre to post-exercise in all participants (n = 13). P placebo; Q quercetin; PRE pre-exercise; POS post-exercise; Bonf Bonferroni; log log-transformed

| Glycan Trait | Pair | PRE mean | POS mean | Delta POS minus PRE | t | p raw | p Bonf |
| --- | --- | --- | --- | --- | --- | --- | --- |
| Btotal | Day21-P | 11.7826<br>377 | 11.6682<br>27 | -0.114410769 | 0.262514 | 0.797378 | 1 |
| Btotal | Day21-Q | 12.5027<br>4 | 12.0172<br>18 | -0.485522308 | 1.441841 | 0.174933 | 0.3498655<br>54 |
| Btotal | Day1-P | 12.1972<br>646 | 12.0546<br>22 | -0.142643077 | 0.934569 | 0.368436 | 0.7368719<br>42 |
| Btotal | Day1-Q | 11.8097<br>946 | 11.7471<br>84 | -0.062610769 | 0.582852 | 0.570784 | 1 |
| G0 (log) | Day21-P | 1.81951<br>403 | 1.90144<br>43 | 0.081930246 | -1.72225 | 0.110673 | 0.2213454<br>06 |
| G0 (log) | Day21-Q | 1.94577<br>222 | 1.94344<br>15 | -0.002330727 | 0.11765 | 0.908292 | 1 |
| **G0 (log)** | **Day1-P** | **1.94849**<br>**571** | **1.91117**<br>**34** | **-0.03732229** | **2.677706** | **0.020122** | **0.0402441**<br>**81** |
| G0 (log) | Day1-Q | 1.90651<br>046 | 1.90955<br>2 | 0.003041533 | -0.21467 | 0.833632 | 1 |
| G1 | Day21-P | 9.66685 | 10.3129<br>16 | 0.646066154 | -1.60576 | 0.134304 | 0.2686088<br>4 |
| G1 | Day21-Q | 10.5681<br>185 | 10.6605<br>86 | 0.092467692 | -0.38825 | 0.704632 | 1 |
| G1 | Day1-P | 10.8011<br>369 | 10.4802<br>88 | -0.320849231 | 2.272959 | 0.042212 | 0.0844237<br>19 |
| G1 | Day1-Q | 10.4447<br>092 | 10.4682<br>03 | 0.023493846 | -0.1476 | 0.885108 | 1 |
| G2 | Day21-P | 56.8133<br>785 | 54.9966<br>32 | -1.816746923 | 2.250733 | 0.043939 | 0.0878782<br>23 |
| G2 | Day21-Q | 55.3154<br>885 | 55.2787<br>9 | -0.036698462 | 0.064035 | 0.949997 | 1 |
| **G2** | **Day1-P** | **54.7498**<br>**7** | **55.8448**<br>**88** | **1.095017692** | **-4.07702** | **0.001534** | **0.0030683**<br>**53** |
| G2 | Day1-Q | 55.1573<br>838 | 55.0712<br>35 | -0.086148462 | 0.266198 | 0.794606 | 1 |
| Gtotal | Day21-P | 86.7290<br>931 | 86.1032<br>65 | -0.625828462 | 1.861327 | 0.087363 | 0.1747259<br>93 |
| Gtotal | Day21-Q | 85.9912<br>708 | 85.9838<br>24 | -0.007446923 | 0.040886 | 0.968059 | 1 |
| **Gtotal** | **Day1-P** | **85.9320**<br>**231** | **86.2489**<br>**42** | **0.316919231** | **-3.1114** | **0.008997** | **0.0179948**<br>**01** |
| Gtotal | Day1-Q | 86.1866<br>1 | 86.1220<br>56 | -0.064553846 | 0.532988 | 0.603773 | 1 |
| Stotal | Day21-P | 72.6244<br>623 | 71.2029<br>15 | -1.421546923 | 1.690574 | 0.116704 | 0.2334085<br>64 |
| Stotal | Day21-Q | 70.5947<br>631 | 70.5400<br>32 | -0.054730769 | 0.120362 | 0.906188 | 1 |
| Stotal | Day1-P | 70.3690<br>654 | 70.9691<br>18 | 0.600052308 | -2.40729 | 0.033076 | 0.0661519<br>75 |
| Stotal | Day1-Q | 71.0644<br>254 | 70.9953<br>62 | -0.069063846 | 0.238762 | 0.815319 | 1 |

